# External validation of the EK-LEF electrocardiogram model for left ventricular systolic dysfunction in three U.S. health systems

**DOI:** 10.64898/2026.11.20.26368604

**Authors:** J. Malinkovich, M. Leshchiner, B. Knight, J. Gong, T. Sinha

## Abstract

**Background:** Heart failure is projected to affect more than 11 million U.S. adults by 2050, and reduced left ventricular ejection fraction (LVEF) can be present before symptoms appear. Deep learning models can detect reduced LVEF from the 12-lead electrocardiogram (ECG), but most have been tested only in the institutions that developed them, often after excluding patients with prior heart failure or recordings of poor quality. We evaluated EK-LEF, a self-supervised ECG model fine-tuned on low LVEF detection.

**Methods:** A bidirectional Mamba encoder was pre-trained on 435,420 unlabeled MIMIC-IV ECGs and fine-tuned on 471,248 ECG–echocardiogram pairs from 235,969 patients at Mayo Clinic Rochester and the Mayo Clinic Health System. Mayo Clinic Florida served as the validation set and Mayo Clinic Arizona, a geographically distinct region, as the internal test set, each evaluated with one pair per patient. For external validation, Dandelion Health ran the model on one ECG from each of 17,500 patients at five hospitals in three U.S. health systems. The reference standard was LVEF ≤ 40% on transthoracic echocardiography within 15 days of the ECG. The primary outcomes were sensitivity and specificity.

**Results:** In the Florida validation set of 14,880 patients, 9.9% of whom had LVEF ≤ 40%, the area under the receiver operating characteristic curve (AUROC) was 0.951 (95% CI 0.947–0.955). In the Arizona internal test set of 16,298 patients (prevalence 9.2%), it was 0.955 (0.950–0.959), with a sensitivity of 85.0% (83.1–86.8) and a specificity of 91.7% (91.3–92.2). The signal-quality rule excluded 2.2% of internal test recordings and 0.9% of external ECGs. Among the 17,344 external patients, of whom 9.4% had LVEF ≤ 40%, the model reached an AUROC of 0.912 (0.906–0.919), a sensitivity of 85.7% (84.1–87.4) and a specificity of 81.8% (81.3–82.4), and its negative predictive value was 98.2% (98.0–98.5). Sensitivity ranged from 79.5% to 90.2% between hospitals, and specificity was lowest in patients with a recorded cardiomyopathy (56.1%).

**Conclusions:** EK-LEF retained high sensitivity and specificity at hospitals outside the health system in which it was developed. A negative result may help identify patients who do not need echocardiography.

## Introduction

In the United States, about 6.7 million people over 20 years of age have heart failure, and this number is projected to rise to 11.4 million by 2050[1]. Heart failure with reduced left ventricular ejection fraction (LVEF) is usually diagnosed after symptoms have appeared. It is preceded by a period of asymptomatic left ventricular systolic dysfunction. This affects 3% to 6% of adults in community studies and carries an increased risk of heart failure and death [2]. Treatment at this stage is effective: in the SOLVD prevention trial, enalapril reduced the development of heart failure in patients with a low ejection fraction and no symptoms [3]. Current guidelines recommend treating these patients [4].

However, many of these patients are never identified, as echocardiography is not a routine test in people without symptoms. Guidelines favor natriuretic peptide testing followed by echocardiography in those with raised values. This strategy was tested in selected high-risk patients and requires a blood test for every person screened and a referral for those with raised values [5].

The 12-lead ECG is inexpensive, widely available in primary care and in hospitals, and carries information about ventricular function that a trained neural network can extract [6]. Deep learning models trained on paired ECGs and echocardiograms have detected reduced LVEF with high accuracy in their development cohorts [7–9], including a community cohort in which 2% of participants were affected [10]. In a pragmatic primary care trial, giving clinicians access to such a model increased new diagnoses of low ejection fraction [11]. However, most of these models were evaluated within the institutions that developed them [8]. When they have been applied elsewhere, the area under the receiver operating characteristic curve (AUROC) has fallen by as much as 0.07 [12], although one multisite study found that performance was maintained across four U.S. sites [13]. Because such evidence is still scarce, external testing remains an important step before a model is used in routine care.

We developed the Electrokare Low Ejection Fraction algorithm (EK-LEF) using ECG and echocardiography data from Mayo Clinic. EK-LEF was first pre-trained on unlabeled ECGs from the public MIMIC-IV-ECG database [14], an approach that has improved label efficiency in other ECG models [15,16], and then fine-tuned on Mayo data, where its cut-off was chosen. In this study, we evaluated EK-LEF at five hospitals in three other U.S. health systems. We also examined accuracy by hospital and by patient group. Accuracy by race and ethnicity has been reported within the Mayo Clinic health system [17] but rarely in external cohorts.

## Methods

### Study design and data sources

This was a retrospective study of diagnostic accuracy, and all data were de-identified before we received them. Under the U.S. Common Rule (45 CFR 46) the work did not involve human subjects and did not require institutional review board review or informed consent. The model was developed with ECG–echocardiogram pairs from Mayo Clinic: pairs from Mayo Clinic Rochester and the Mayo Clinic Health System (MCHS) were used for training, pairs from Mayo Clinic Florida for early stopping and model selection, and pairs from Mayo Clinic Arizona, a geographically distinct census region held out from development, as the test set. We assigned each patient to a single Mayo cohort, and the 10,591 patients with records at both Rochester and MCHS were assigned to the training set. For the external test, we licensed de-identified data from Dandelion Health covering five hospitals (Sites 1–5) in Sanford Health, Sharp HealthCare and Texas Health Resources.

For the Mayo cohorts, a pair consisted of an adult 12-lead ECG (10 s, 500 Hz, all leads present) and a quantitative LVEF measured at the same site. We preferred an ECG recorded in the 15 days before the echocardiogram. If none was available, an ECG from the 15 days after it was accepted when LVEF was above 40%. When several ECGs qualified, the one closest in time was chosen. Patients with a previous low LVEF were not excluded, and a patient could contribute more than one pair (Fig. 1).

**Figure 1.**
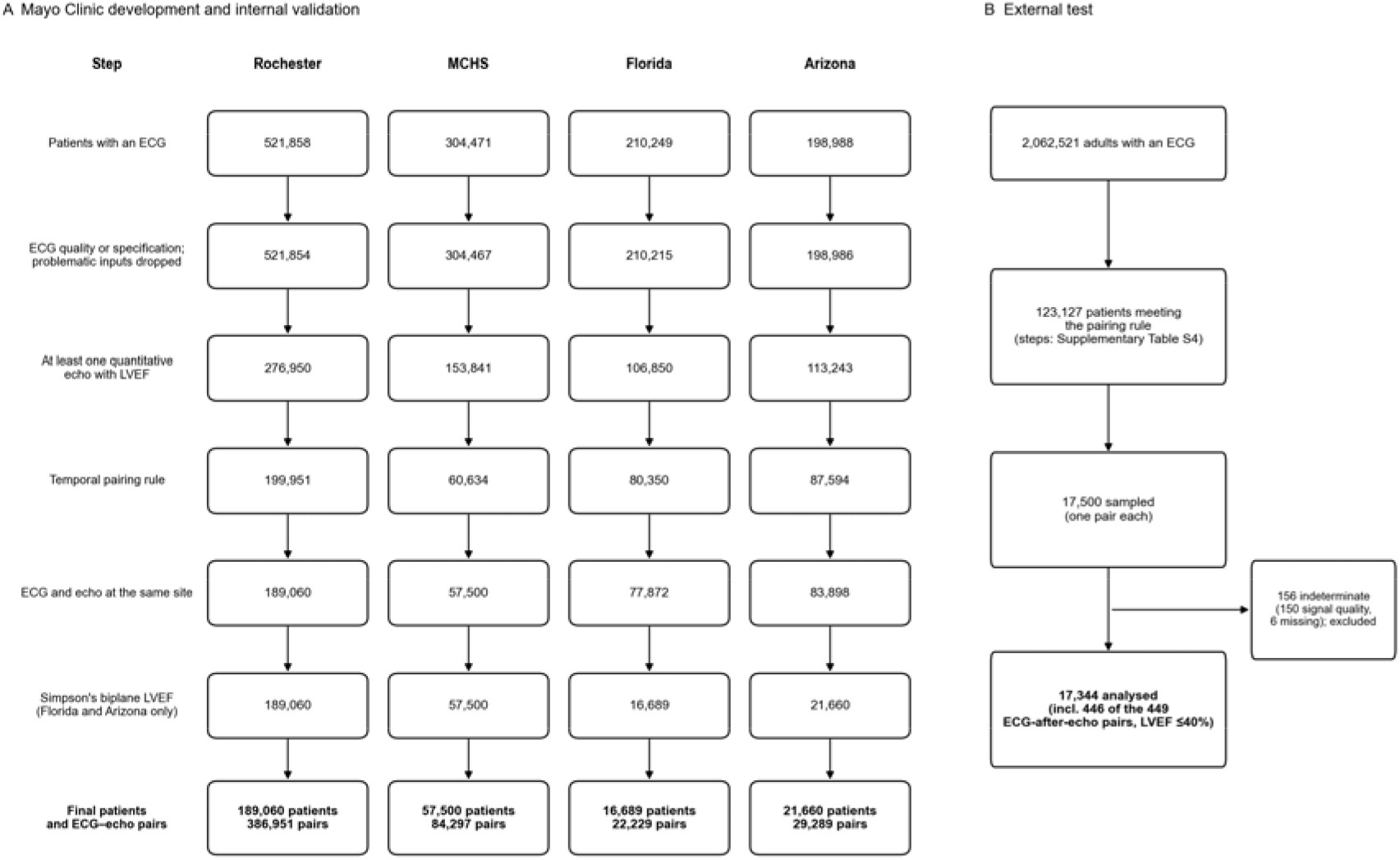
Participant flow for the Mayo Clinic cohorts and the external test, with the number of patients remaining after each screening step. ECG, electrocardiogram; LVEF, left ventricular ejection fraction; MCHS, Mayo Clinic Health System.

Dandelion Health assembled the external cohort from its de-identified database, which links ECG waveforms from the MUSE cardiology information system (GE HealthCare, Chicago, IL, USA) with echocardiography reports and electronic health record data. Adults were eligible if they had a non-paced 10-s 12-lead ECG sampled at 500 Hz or more and a transthoracic Simpson’s biplane LVEF, of any value, within 15 days before or after it. From each hospital, 3,500 eligible patients were sampled at random, and for each patient the ECG–echocardiogram pair with the shortest interval was selected. The index ECGs were recorded between June 2016 and April 2026, 69% of them during inpatient encounters. Dandelion Health ran the model on its own platform, storing the outputs before the echocardiographic data were linked, and returned only aggregate results to us.

### Reference standard and quality screen

The reference standard was an LVEF of 40% or less on transthoracic echocardiography, the threshold used to define heart failure with reduced ejection fraction [4,18]. Only Simpson’s biplane measurements were accepted at Florida, Arizona and the external sites. Rochester and MCHS accepted LVEF measured by any quantitative method. The software’s built-in quality check withholds a result when the signal-to-noise ratio is below 5.0 dB in lead II and in any of V2–V4. External ECGs that failed this check were recorded as indeterminate.

### Model development

EK-LEF reads the eight independent leads (I, II and V1–V6) of a 10-s ECG sampled at 500 Hz, after each lead has been normalized in amplitude; preprocessing was the same at every site. A convolutional tokenizer divides each lead into 416 tokens, and a six-layer bidirectional Mamba encoder then processes all 3,328 tokens as a single sequence (Fig. 2). Mamba is a state-space model whose computational cost grows linearly with sequence length [19]. State-space models have performed well in ECG classification [20], and a bidirectional encoder processes each segment of the trace together with the signal before and after it, as in earlier bidirectional state-space models [21,22]. The encoder was pre-trained on 435,420 recordings from MIMIC-IV-ECG [14] with a joint-embedding predictive architecture (JEPA) [23], learning to predict the embeddings of masked segments from the visible parts of the same and other leads. In half of the pre-training ECGs, a random subset of leads was dropped and replaced by a learned absent-lead embedding. Pre-training used AdamW with a learning rate of 5 × 10⁻⁴ and a batch size of 64.

**Figure 2.**
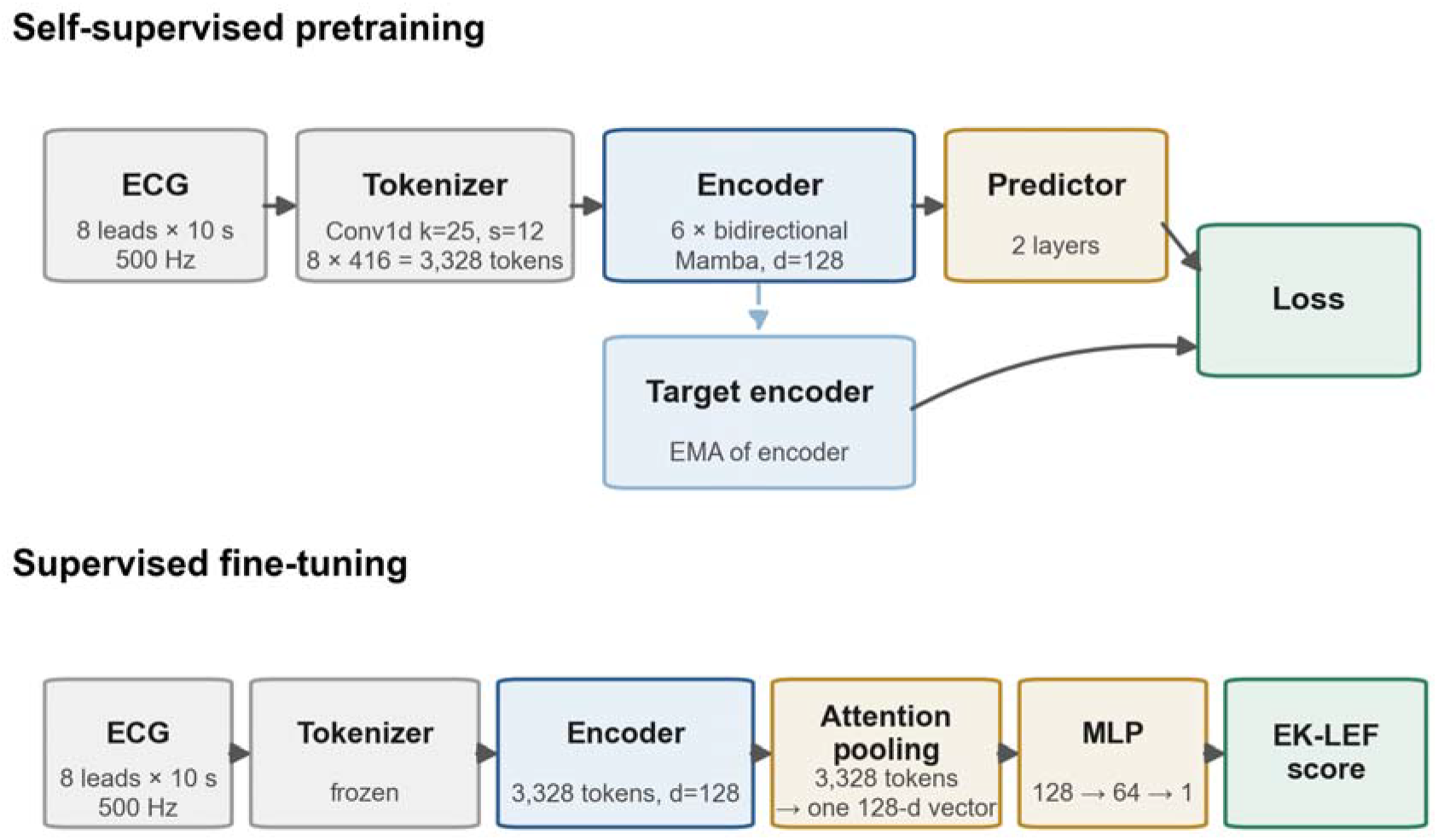
EK-LEF architecture. Top, JEPA pre-training with a two-layer predictor and a target encoder updated as an exponential moving average (EMA) of the encoder. Bottom, supervised fine-tuning with a frozen tokenizer, attention pooling and a multilayer perceptron (MLP) head. ECG, electrocardiogram; JEPA, joint-embedding predictive architecture.

For fine-tuning, we froze the tokenizer and trained the rest of the network on 471,248 ECG–echocardiogram pairs from 235,969 Rochester and MCHS patients, stopping early on the Florida pairs with a patience of 4 epochs. Fine-tuning converged within 6 epochs, and the model from epoch 6 was used for all analyses. The loss was a class-weighted focal binary cross-entropy, optimized with AdamW at learning rates of 5 × 10⁻⁵ for the output head and 2.5 × 10⁻⁵ for the encoder, with a batch size of 256. The operating threshold, 0.350, was the unweighted mean of the optimal thresholds in seven development units (Rochester, five MCHS sites and Florida), found by F1 score at Rochester and Florida and by Youden’s J at the MCHS sites. This threshold was then applied to Arizona and to the external sites.

### Statistical analysis

Accuracy was calculated per ECG–echocardiogram pair. The primary analysis tested whether the lower 95% confidence limits of external sensitivity and specificity were both at least 80%. With 3,500 patients per hospital, accuracy could also be examined by site, sex, race, ethnicity, age, comorbidity and ECG device. For comparison with the external cohort, Florida and Arizona were also evaluated with exactly one pair per patient, selecting the ECG and echocardiogram closest in time within 15 days before or after and excluding ECGs in which every lead was flat after the first 5 s. After the quality rule, which removed 1.6% and 2.2% of the remaining recordings, the sets included 14,880 and 16,298 patients. The report follows STARD 2015 [24]. Definitions, training settings, the derivation of the threshold and changes to the study plan are given in the Supplementary Appendix, and the completed STARD checklist is provided as a separate file.

Continuous variables are given as mean (SD) or median, and discrimination as the AUROC and the average precision. For the external cohort, 95% CIs were obtained by percentile bootstrap over patients (2,000 replicates); for Arizona, Wilson intervals over pairs were used for sensitivity and specificity. Indeterminate outputs were excluded from the primary analysis. In addition, in sensitivity analyses they were counted first as correct and then as incorrect, and pairs were also restricted by the timing of the ECG relative to the echocardiogram. Sites and subgroups were compared with unadjusted χ² tests, without correction for multiple comparisons. A two-sided P value below 0.05 was considered statistically significant. We also examined test positivity by LVEF band, with Clopper–Pearson intervals, and calibration by score decile. Analyses were done in Python (Python Software Foundation) with pandas, NumPy and SciPy, and in PyTorch (PyTorch Foundation).

## Results

### Participants

Table 1 describes the cohorts. External patients had a mean age of 65.5 years (SD 15.7) and a mean LVEF of 58.1%, and the median interval between ECG and echocardiogram was 3.4 h, with 78% of pairs recorded within 24 h of each other. Compared with the training cohorts, the external cohort included more women (48.1% vs 43.1% at Rochester and 44.9% at MCHS) and more Black patients (7.2% vs 2.2% and 1.6%). Of the 17,500 external patients, 156 had no valid output, leaving 17,344 for analysis, of whom 1,625 (9.4%) had LVEF ≤ 40% (Fig. 1).

**Table 1.** Characteristics of the development, internal and external cohorts.

| **Characteristic** | **Rochester** | **MCHS** | **Florida** | **Arizona** | **External** |
|---|---|---|---|---|---|
| Patients, n | 189,060 | 57,500 | 16,689 | 21,660 | 17,500 |
| ECG–echo pairs, n | 386,951 | 84,297 | 22,229 | 29,289 | 17,500 |
| LVEF ≤ 40%, n (%) | 20,937 (5.4%) | 6,795 (8.1%) | 1,299 (5.8%) | 1,723 (5.9%) | 1,633 (9.3%) |
| Age, years, mean (SD) | 60.5 (15.5) | 65.6 (14.5) | 63.2 (14.7) | 64.3 (14.3) | 65.5 (15.7) |
| Male, n (%) | 220,284 (56.9%) | 46,410 (55.1%) | 12,528 (56.4%) | 18,547 (63.3%) | 9,084 (51.9%) |
| Female, n (%) | 166,652 (43.1%) | 37,887 (44.9%) | 9,701 (43.6%) | 10,735 (36.7%) | 8,415 (48.1%) |
| White, n (%) | 357,546 (92.4%) | 81,187 (96.3%) | 18,599 (83.7%) | 25,883 (88.4%) | 13,618 (77.8%) |
| Black or African American, n (%) | 8,352 (2.2%) | 1,313 (1.6%) | 2,432 (10.9%) | 1,042 (3.6%) | 1,258 (7.2%) |
| Hispanic or Latino, n (%) | 8,438 (2.2%) | 1,544 (1.8%) | 1,072 (4.8%) | 2,422 (8.3%) | 1,610 (9.2%) |

For the Mayo cohorts, percentages refer to ECG–echocardiogram pairs; for the external cohort, they refer to patients. Further characteristics and the accounting of the external cohort are given in the Supplementary Appendix. ECG, electrocardiogram; echo, echocardiogram; LVEF, left ventricular ejection fraction; MCHS, Mayo Clinic Health System; SD, standard deviation.

External CIs: patient-level percentile bootstrap (2,000 resamples). Sites are numbered in order of prevalence; Sites 2 and 3 are two different Sanford Health hospitals, and Sites 4 and 5 two different Texas Health Resources hospitals. The 156 indeterminate outputs are excluded from the primary and site rows; in the best and worst cases they are counted as correct and as incorrect, with a score of 1 or 0 for AUROC. Arizona CIs: Wilson intervals, and for AUROC a percentile bootstrap (2,000 resamples). Development cohorts are not shown. AUROC, area under the receiver operating characteristic curve; CI, confidence interval; ECG, electrocardiogram; echo, echocardiogram; FN, false negatives; FP, false positives; LVEF, left ventricular ejection fraction; NPV, negative predictive value; PPV, positive predictive value; TN, true negatives; TP, true positives.

### External test

In the external cohort, EK-LEF had an AUROC of 0.912 (95% CI 0.906–0.919) and an average precision of 0.579. At the operating threshold of 0.350, its sensitivity was 85.7% (84.1–87.4) and its specificity was 81.8% (81.3–82.4), with 1,393 true-positive and 2,856 false-positive results. Table 2 shows these results by hospital, and Fig. 3 shows the confusion matrices. The PPV was 32.8%, and the NPV was 98.2%. In Arizona, the AUROC was 0.955 (0.950–0.959) with one pair per patient. At the operating threshold, sensitivity was 85.0% (83.1–86.8) and specificity 91.7% (91.3–92.2), with a PPV of 51.0% and an NPV of 98.4%. This set included 16,298 patients with a mean age of 63.8 years (SD 14.8), of whom 61.3% were male and 1,497 (9.2%) had LVEF ≤ 40%. The AUROC was 0.929 (0.923–0.934) across all 29,289 pairs, in which sensitivity was 83.3% (81.5–85.0) and specificity 86.2% (85.8–86.6) at the operating threshold. In the Florida validation set, which was used for early stopping and threshold selection, the AUROC was 0.951 (0.947–0.955) in 14,880 patients, of whom 1,479 (9.9%) had LVEF ≤ 40%; sensitivity was 86.1% (84.3–87.8) and specificity 89.0% (88.5–89.6).

**Figure 3.**
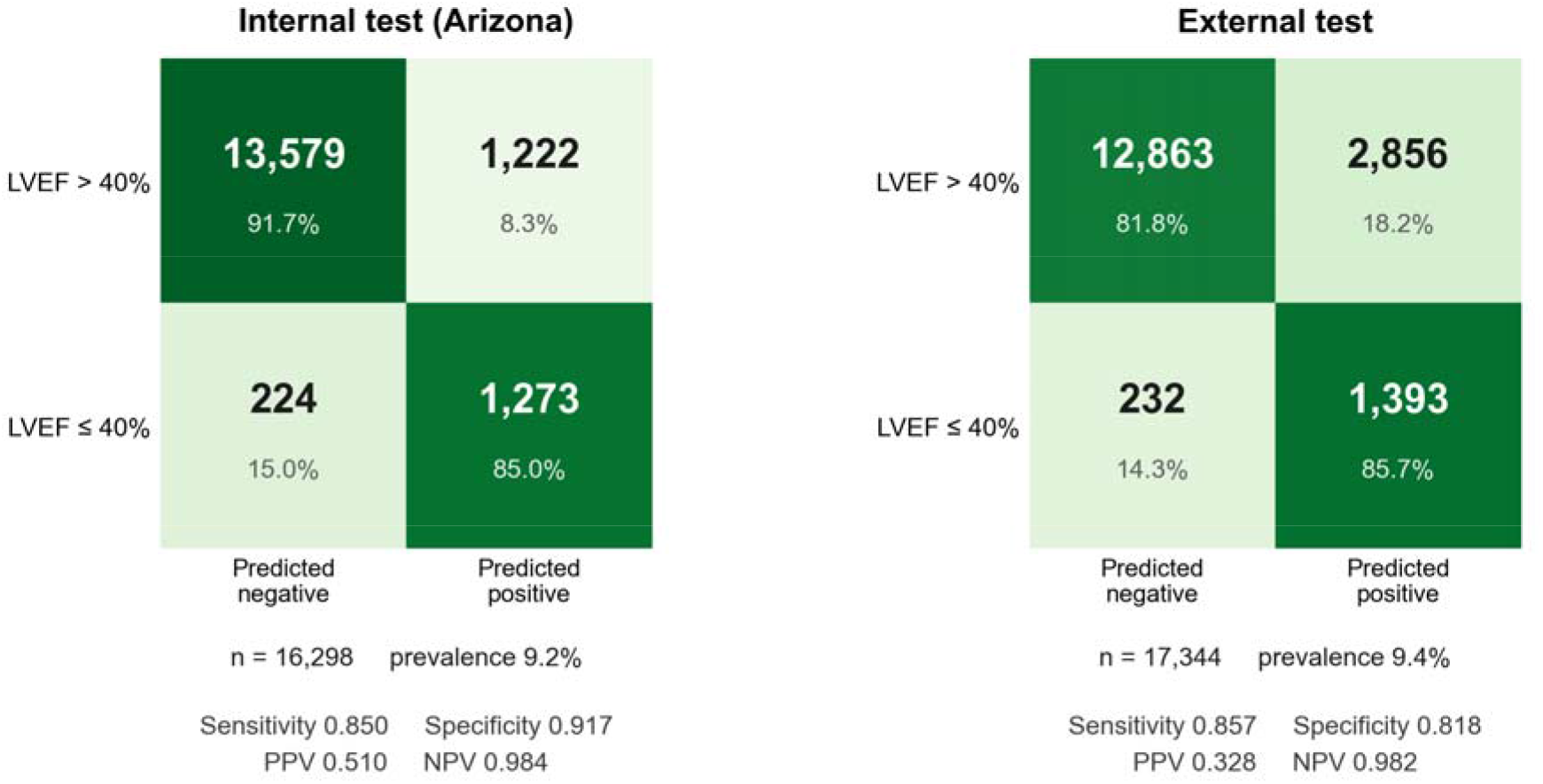
Confusion matrices at the operating threshold for Arizona (internal test set) and the external cohort, each with one pair per patient. Row percentages are shown. LVEF, left ventricular ejection fraction; NPV, negative predictive value; PPV, positive predictive value.

**Table 2.**
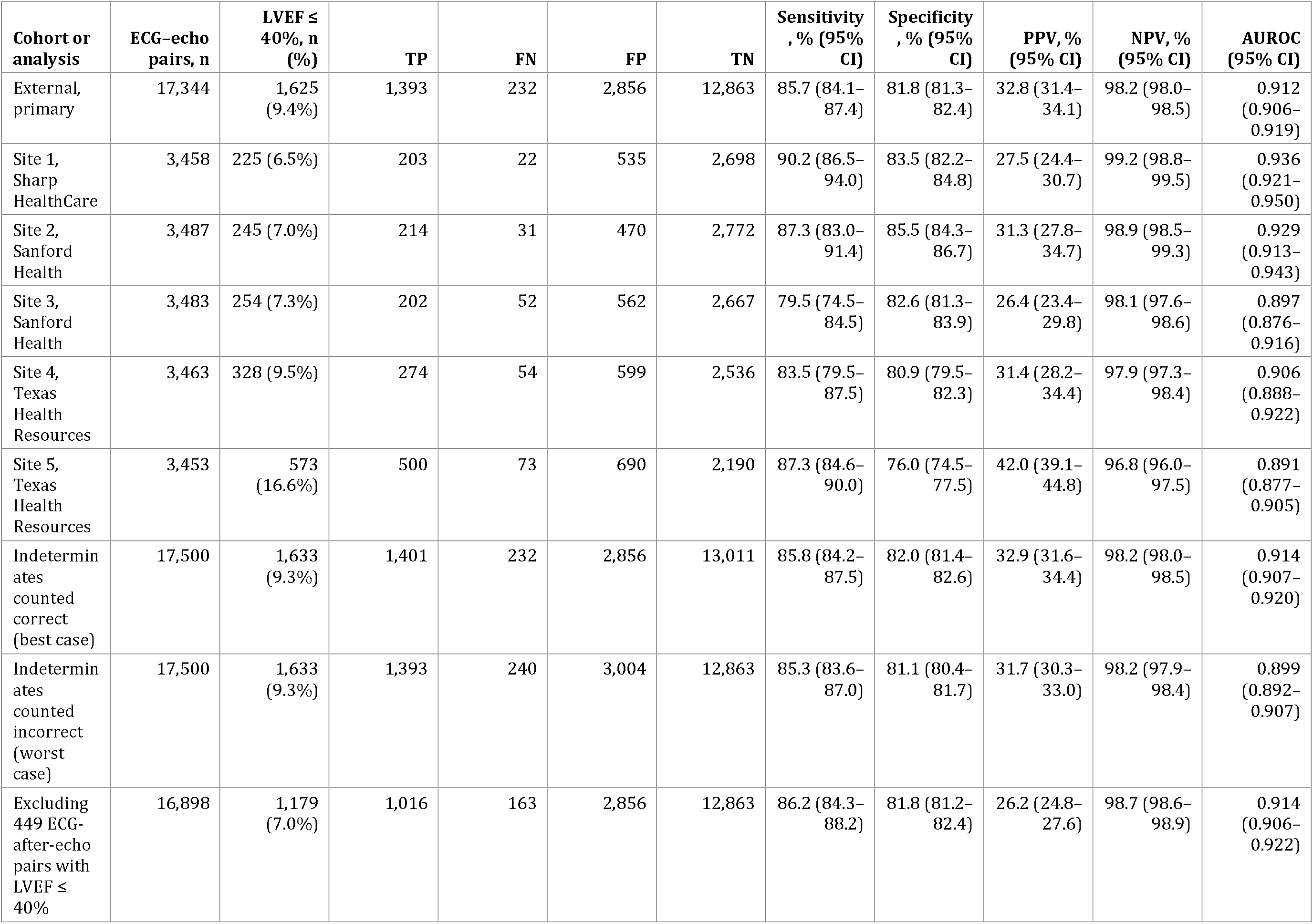
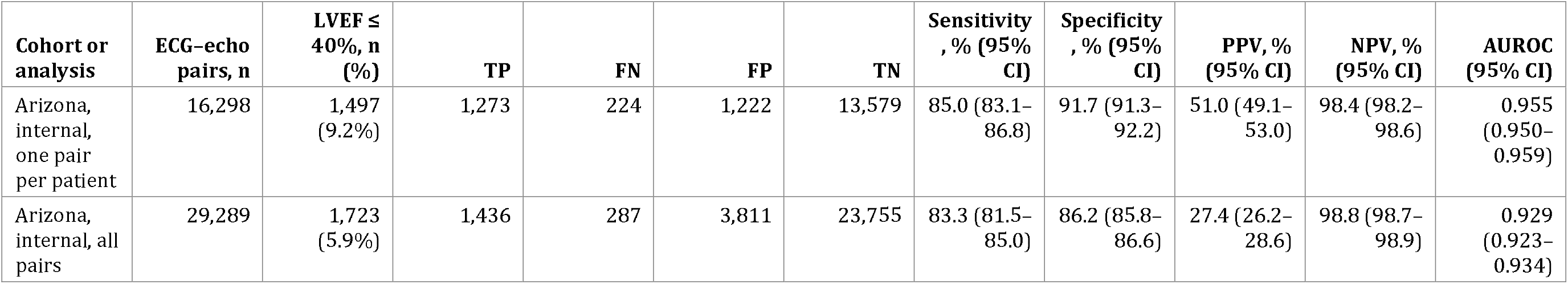
Accuracy in the external cohort (primary) and Arizona (internal, secondary)

| Cohort or analysis | ECG–echo pairs, n | LVEF ≤ 40%, n (%) | TP | FN | FP | TN | Sensitivity, % (95% CI) | Specificity, % (95% CI) | PPV, % (95% CI) | NPV, % (95% CI) | AUROC (95% CI) |
|---|---|---|---|---|---|---|---|---|---|---|---|
| External, primary | 17,344 | 1,625 (9.4%) | 1,393 | 232 | 2,856 | 12,863 | 85.7 (84.1–87.4) | 81.8 (81.3–82.4) | 32.8 (31.4–34.1) | 98.2 (98.0–98.5) | 0.912 (0.906–0.919) |
| Site 1, Sharp HealthCare | 3,458 | 225 (6.5%) | 203 | 22 | 535 | 2,698 | 90.2 (86.5–94.0) | 83.5 (82.2–84.8) | 27.5 (24.4–30.7) | 99.2 (98.8–99.5) | 0.936 (0.921–0.950) |
| Site 2, Sanford Health | 3,487 | 245 (7.0%) | 214 | 31 | 470 | 2,772 | 87.3 (83.0–91.4) | 85.5 (84.3–86.7) | 31.3 (27.8–34.7) | 98.9 (98.5–99.3) | 0.929 (0.913–0.943) |
| Site 3, Sanford Health | 3,483 | 254 (7.3%) | 202 | 52 | 562 | 2,667 | 79.5 (74.5–84.5) | 82.6 (81.3–83.9) | 26.4 (23.4–29.8) | 98.1 (97.6–98.6) | 0.897 (0.876–0.916) |
| Site 4, Texas Health Resources | 3,463 | 328 (9.5%) | 274 | 54 | 599 | 2,536 | 83.5 (79.5–87.5) | 80.9 (79.5–82.3) | 31.4 (28.2–34.4) | 97.9 (97.3–98.4) | 0.906 (0.888–0.922) |
| Site 5, Texas Health Resources | 3,453 | 573 (16.6%) | 500 | 73 | 690 | 2,190 | 87.3 (84.6–90.0) | 76.0 (74.5–77.5) | 42.0 (39.1–44.8) | 96.8 (96.0–97.5) | 0.891 (0.877–0.905) |
| Indeterminates counted correct (best case) | 17,500 | 1,633 (9.3%) | 1,401 | 232 | 2,856 | 13,011 | 85.8 (84.2–87.5) | 82.0 (81.4–82.6) | 32.9 (31.6–34.4) | 98.2 (98.0–98.5) | 0.914 (0.907–0.920) |
| Indeterminates counted incorrect (worst case) | 17,500 | 1,633 (9.3%) | 1,393 | 240 | 3,004 | 12,863 | 85.3 (83.6–87.0) | 81.1 (80.4–81.7) | 31.7 (30.3–33.0) | 98.2 (97.9–98.4) | 0.899 (0.892–0.907) |
| Excluding 449 ECG-after-echo pairs with LVEF ≤ 40% | 16,898 | 1,179 (7.0%) | 1,016 | 163 | 2,856 | 12,863 | 86.2 (84.3–88.2) | 81.8 (81.2–82.4) | 26.2 (24.8–27.6) | 98.7 (98.6–98.9) | 0.914 (0.906–0.922) |
| **Cohort or analysis** | **ECG–echo pairs, n** | **LVEF ≤ 40%, n (%)** | **TP** | **FN** | **FP** | **TN** | **Sensitivity, % (95% CI)** | **Specificity, % (95% CI)** | **PPV, % (95% CI)** | **NPV, % (95% CI)** | **AUROC (95% CI)** |
| Arizona, internal, one pair per patient | 16,298 | 1,497 (9.2%) | 1,273 | 224 | 1,222 | 13,579 | 85.0 (83.1–86.8) | 91.7 (91.3–92.2) | 51.0 (49.1–53.0) | 98.4 (98.2–98.6) | 0.955 (0.950–0.959) |
| Arizona, internal, all pairs | 29,289 | 1,723 (5.9%) | 1,436 | 287 | 3,811 | 23,755 | 83.3 (81.5–85.0) | 86.2 (85.8–86.6) | 27.4 (26.2–28.6) | 98.8 (98.7–98.9) | 0.929 (0.923–0.934) |

Sensitivity and specificity both differed significantly between hospitals (P = 0.006 and P < 0.001; Table 2). The hospitals also differed in patient mix and in the ECG equipment they used. Inpatient encounters accounted for 55% of index ECGs at Site 2 and 83% at Site 5. The two Sanford Health hospitals (Sites 2 and 3) recorded most ECGs on Philips PageWriter TC70 machines. The other three hospitals used GE devices. Sensitivity was highest at Site 1 (90.2%), and Site 5, which had the most inpatients and the highest prevalence (16.6%), had the lowest specificity (76.0%).

### Subgroups and devices

Sensitivity was similar across demographic subgroups, but specificity differed significantly by sex, age and comorbidity (all P < 0.01; Fig. 4A and Supplementary Table S6). Specificity was 84.5% (95% CI 83.7–85.4) in women and 79.2% (78.3–80.1) in men, 82.3% (81.7–83.0) in White and 74.7% (72.0–77.2) in Black patients, and 77.8% (75.7–79.8) in patients aged 85 years or older. It was lowest in patients with a recorded cardiomyopathy (56.1%, 53.0–59.0) or end-stage renal disease (63.4%, 59.6–67.2). Specificity was also low in patients with persistent or chronic atrial fibrillation (66.5%, 64.0–69.2) or a heart failure diagnosis (67.5%, 66.0–68.9). Specificity was 79.8% for ECGs acquired on GE MAC 5500 machines and 84.9% for those acquired on Philips PageWriter TC70 machines.

**Figure 4.**
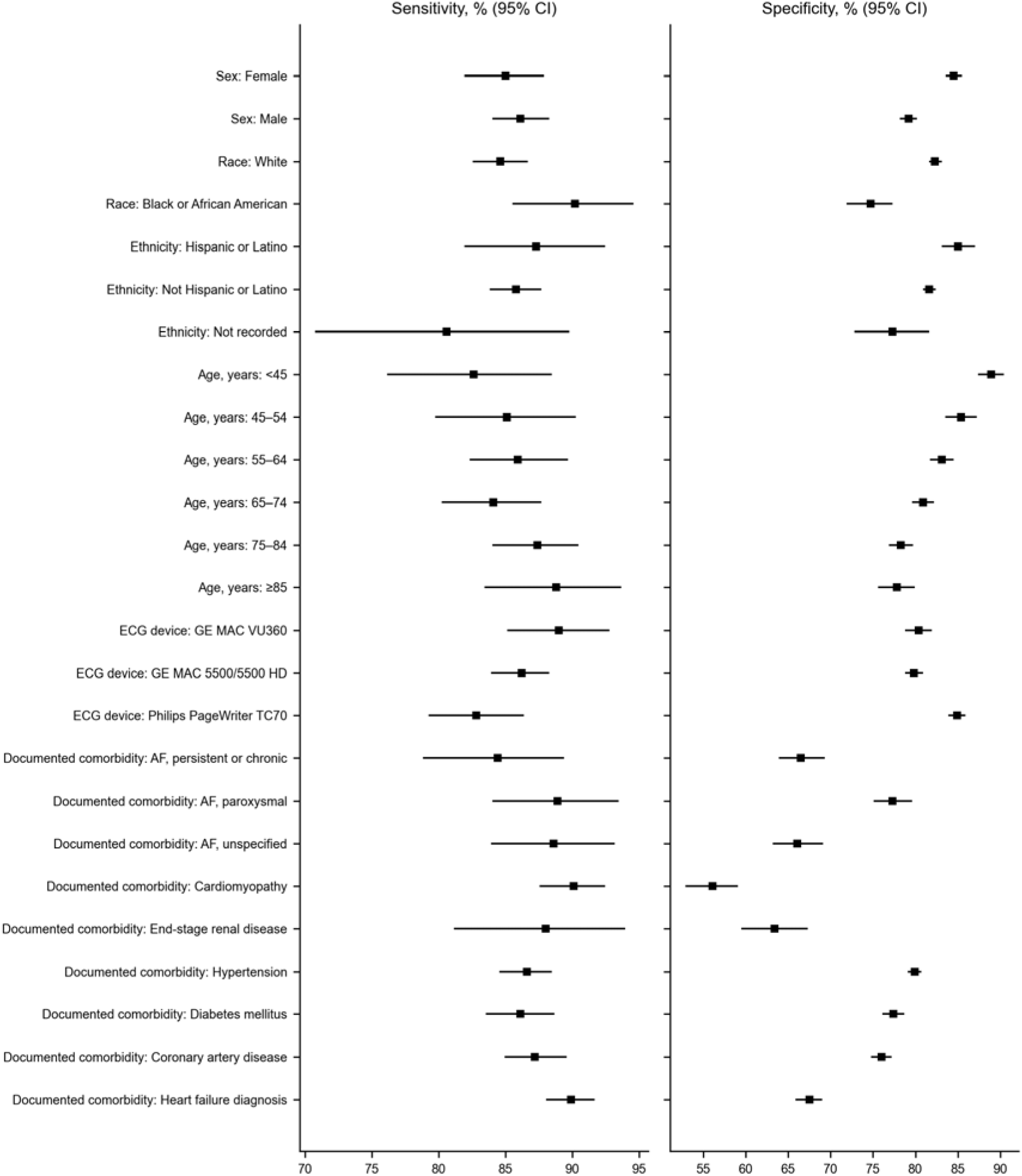
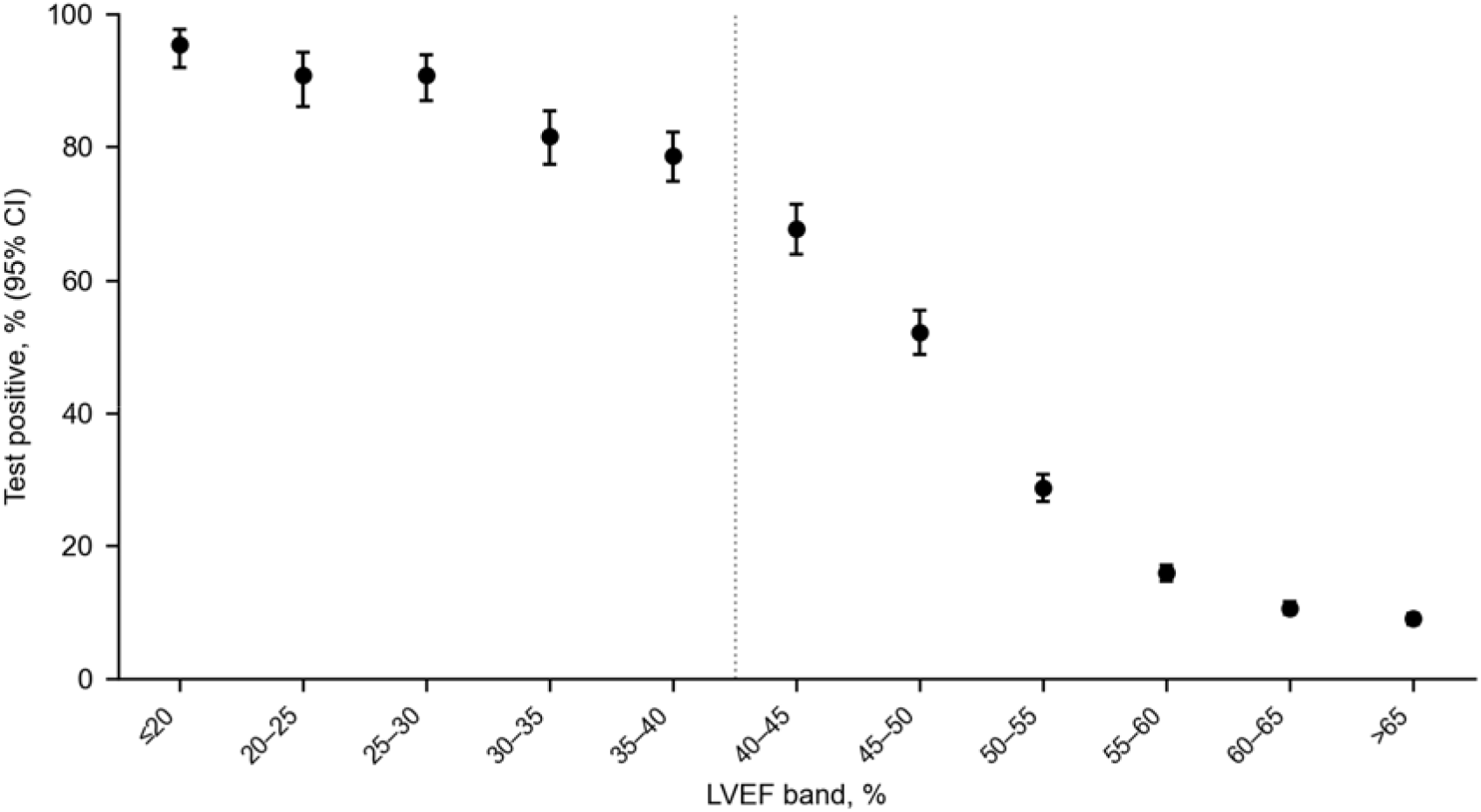
External results by subgroup and by LVEF band, 17,344 patients. (A) Sensitivity and specificity with 95% CIs in each subgroup; strata with a cell below 11 are not shown. (B) Proportion testing positive at the operating threshold in each LVEF band, with exact 95% CIs. CI, confidence interval; LVEF, left ventricular ejection fraction.

### LVEF bands and calibration

Test positivity rose as LVEF fell (Fig. 4B), from 52.1% at LVEF 46–50% and 67.8% at 41–45% to 78.7% at 36–40% and 95.4% at 20% or less. Of the 2,856 false-positive results, 892 (31.2%) were in patients with LVEF 41–50%. The score was poorly calibrated at the low end: in the lowest 30% of scores, the mean score was 0.066 against an observed prevalence of 0.3%, and the calibration slope was 2.18 (Supplementary Table S7).

### Sensitivity analyses

Excluding the 449 pairs in which the ECG followed an echocardiogram showing LVEF ≤ 40% gave a sensitivity and specificity of 86.2% and 81.8%, respectively (Table 2). We found similar results when only ECGs recorded within 1 day of the echocardiogram (sensitivity 87.8%, specificity 81.0%) or before it (86.2% and 82.0%) were included (Supplementary Table S8).

## Discussion

We validated EK-LEF, a self-supervised deep learning ECG algorithm developed with Mayo Clinic data, in three U.S. health systems outside Mayo Clinic, using its original operating threshold. To our knowledge, this is the first multisite external validation of a self-supervised state-space ECG model for LVEF ≤ 40%. At the external hospitals, sensitivity (85.7%) was close to that in Arizona (85.0%), but specificity was lower (81.8% vs 91.7%). With one pair per patient in both settings, the AUROC fell from 0.95 to 0.91. These results are encouraging for clinical use because clinicians act on a binary result, and most hospitals lack the labeled data and the staff to derive a threshold of their own. The decrease we observed is also smaller than that reported when AI-ECG models were moved between institutions in a previous study [12], and it is consistent with the external results of Carter et al. [13] (Supplementary Table S9). Earlier detection of systolic dysfunction could, therefore, shorten the delay to treatment for a common condition that carries a high burden of hospital admission and death [2].

Specificity was the weaker of the two measures, but false positives were concentrated in identifiable groups. Nearly a third of them (892) occurred in patients with LVEF between 41% and 50%, a range that largely overlaps mildly reduced ejection fraction (41–49%) [18], and patients with a recorded cardiomyopathy, end-stage renal disease, atrial fibrillation or heart failure had the lowest specificity. Reassuringly, echocardiography is clinically reasonable for many of these patients, so a positive result may seldom lead to an unnecessary test. In addition, among the hospitals, specificity was highest at the site with the smallest share of inpatients (Site 2) and lowest at the site with the largest (Site 5); it also differed between ECG machine types, although each machine type was confined to particular hospitals. The lower specificity we observed in Black patients may reflect factors that our data could not capture, and it should be examined in a study with access to individual records.

The score overestimated risk at low values and is intended to be reported as a binary result. In this referred population, of whom 9.4% had LVEF ≤ 40%, a negative result had an NPV of 98.2%; among unselected community adults, where prevalence is much lower [10], the NPV would be higher still, while most positive results would be false positives. The algorithm is, therefore, best suited to identifying patients who do not need echocardiography. Moreover, the quality rule withheld a result for 0.9% of external ECGs and 2.2% of internal test recordings, fewer than in several earlier evaluations. Studies of an ECG-enabled stethoscope, a smartwatch and a four-site 12-lead validation excluded between 12.8% and 21.5% of patients or recordings because of poor signal quality or rhythm [13,25,26]. A pragmatic trial of AI-ECG screening also enrolled only adults without prior heart failure [11]. Our external cohort excluded no patients because of prior heart failure or any other diagnosis.

Self-supervised ECG models such as HeartBEiT and ECG-FM have been evaluated on held-out data from their own networks [27,28]. ECG-Fyler, pre-trained mainly on children, showed a marked drop in accuracy in adults at another center [29]. EK-LEF was pre-trained and tested on adults, which is one possible reason for its better external performance. As with other deep learning ECG models, the waveform features that drive the EK-LEF score remain to be established, and future work should examine them with saliency maps.

Our study has several limitations. First, every patient we studied had been referred for echocardiography. These patients have more heart disease than a screening population, and specificity and PPV are likely to differ in screening use. Although a referred population is the usual setting for validation studies of this kind, a prospective evaluation in primary care is needed before such use. Second, the contribution of self-supervised pre-training cannot be measured here, because no comparable model was trained without it. Third, lead dropout during pre-training was meant to let the model work with any subset of leads, but we tested only complete 12-lead ECGs. Fourth, echocardiographic LVEF near 40% is subject to measurement error from apical foreshortening and endocardial dropout [30]. Fifth, as we received only aggregate results, the effects of patient mix and ECG equipment on the differences between sites could not be distinguished, and individual false-positive results were not available for review. Finally, the intervals for the all-pairs Arizona analysis may understate uncertainty, because some patients contributed more than one ECG.

## Conclusions

EK-LEF retained high sensitivity and specificity at five hospitals in three other U.S. health systems. A prospective study in primary care is needed to test whether a negative result can safely replace echocardiography in patients with no other indication for the test.

## Data availability

The data-use agreements with Mayo Clinic and Dandelion Health do not permit sharing of record-level ECG or echocardiography data. Aggregate results are given in the tables and Supplementary Appendix. MIMIC-IV-ECG is available through PhysioNet (https://physionet.org/content/mimic-iv-ecg/).

## Code availability

Model weights and training code are proprietary to Electrokare and are not publicly available. Protocols and statistical code are available from the corresponding author on reasonable request, subject to the data-use agreements.

## Acknowledgements

We thank Mayo Clinic for the development and internal test data, and Dandelion Health for the external data and for running the external analysis.

## Use of AI tools

A large language model (Claude, Anthropic) was used in preparing the manuscript text.

## Supplementary appendix

**Supplementary Table S1.** Eligibility, pairing, quality and comorbidity definitions.

| Item | Mayo cohorts (Rochester, MCHS, Florida, Arizona) | External cohort |
|---|---|---|
| ECG | Standard resting 12-lead, 500 Hz | 12-lead, ≥500 Hz, 10 s, all leads present; paced rhythm or pacemaker excluded |
| Reference standard | Transthoracic LVEF; Simpson’s biplane only at Florida and Arizona, any acceptable method at Rochester and MCHS | Protocol: transthoracic LVEF by Simpson’s biplane; of 17,500 echocardiograms, 17,455 transthoracic, approach unspecified in 32 and missing in 13, all at Sites 3–5 |
| Pairing window | Same-site ECG within 15 days before the echocardiogram preferred; within 15 days after only if LVEF >40% | Same-site ECG within 15 days before or after, any LVEF |
| Several candidate ECGs | Closest in time, ties to the before-echo ECG; each echocardiogram and each ECG in at most one pair | Smallest ECG–echo interval, before or after |
| Pairs per patient | More than one allowed | One |
| Signal-quality rule | Development data: ECGs failing automated signal-quality screening excluded; thresholds and excluded count not recorded. Florida and Arizona one-pair sets: 5 dB signal-to-noise rule on lead II and V2–V4, removing 1.6% and 2.2% of recordings | 5 dB signal-quality screen on lead II and V2–V4: 150 of 17,494 scored ECGs indeterminate; 6 pairs without output; handling in Methods and Fig. 1 |
| One-pair sets, Florida and Arizona | Patients: 16,689 and 21,660 with development pairs; 14,213 and 15,432 after excluding ECGs in which every lead was flat after the first 5 s; 15,126 and 16,666 after adding patients whose ECG followed an echocardiogram showing LVEF ≤ 40%; 14,880 and 16,298 after the 5 dB rule | — |
| Comorbidity dating | Each comorbidity dated by its first documentation | Code documented on or before the index-ECG date |
| LVEF range cleaning | Not recorded | As for the Mayo cohorts |
| Echocardiogram reader blinding | Not recorded | As for the Mayo cohorts |
| Patient overlap check | Method not recorded | Cross-match with Mayo patients not recorded |
| Treatment between ECG and echocardiogram | Not recorded | Not recorded |
| **External condition** | **Codes and qualifiers** |  |
| Atrial fibrillation | Persistent/chronic I48.1, I48.2, taking precedence; paroxysmal I48.0 without a persistent/chronic code; unspecified I48.9, I48.91 without either; I48.92 (atrial flutter) excluded |  |
| Cardiomyopathy | I42, I43, I25.5, O90.3 |  |
| CKD stage 3–5; ESRD | N18.3–N18.5 (eGFR not used); N18.6 |  |
| Diabetes mellitus | E08–E11, E13 |  |
| Hypertension | I10–I13, I15, I16 |  |
| Coronary artery disease | I20, I25, Z95.1, Z95.5, Z98.61 |  |
| Conduction disorder | I44, I45 |  |
| Heart failure diagnosis | I50, I11.0, I13.0, I13.2 |  |
| History of myocardial infarction | I25.2 at any prior date, or I21/I22 first documented at least 28 days before the index ECG; acute myocardial infarction codes within 28 days do not qualify |  |
| Aortic stenosis, nonrheumatic | I35.0, I35.2 |  |
| Obesity diagnosis | E66, Z68.3, Z68.4 |  |
| Metabolic syndrome | E88.81, E88.810 |  |
| Tobacco use, current or former | F17, Z72.0, Z87.891 |  |
| Alcohol-related disorder or alcoholic liver disease | F10, K70 |  |
| Hyperthyroidism | E05 |  |
| Delivery within 180 days | O80, O82, Z37; latest delivery code within the 180 days ending on the index-ECG date |  |
| Cardiotoxic medication orders | Doxorubicin, daunorubicin, epirubicin, idarubicin, mitoxantrone, trastuzumab, pertuzumab, sunitinib, sorafenib, pazopanib, lapatinib, imatinib, dasatinib, nilotinib, ponatinib, ibrutinib, osimertinib, cyclophosphamide, ifosfamide, carfilzomib, bevacizumab, or ATC class L01DB/L01FD; canceled or deleted orders excluded; order date on or before the index ECG; orders, not administrations |  |

High-risk groups overlap; ≥1, ≥2 and ≥3 count qualifying groups. External, thirteen groups: history of myocardial infarction, cardiomyopathy, nonrheumatic aortic stenosis, persistent/chronic atrial fibrillation, CKD stage 3–5, diabetes mellitus, hypertension, coronary artery disease, conduction disorder, heart failure diagnosis, metabolic syndrome, cardiotoxic medication orders and delivery within 180 days. Methods and Supplementary Table S2 describe the index test. ATC, Anatomical Therapeutic Chemical classification; CKD, chronic kidney disease; ECG, electrocardiogram; echo, echocardiogram; eGFR, estimated glomerular filtration rate; ESRD, end-stage renal disease; ICD-10, International Classification of Diseases, 10th revision; LVEF, left ventricular ejection fraction; MCHS, Mayo Clinic Health System.

**Supplementary Table S2.**
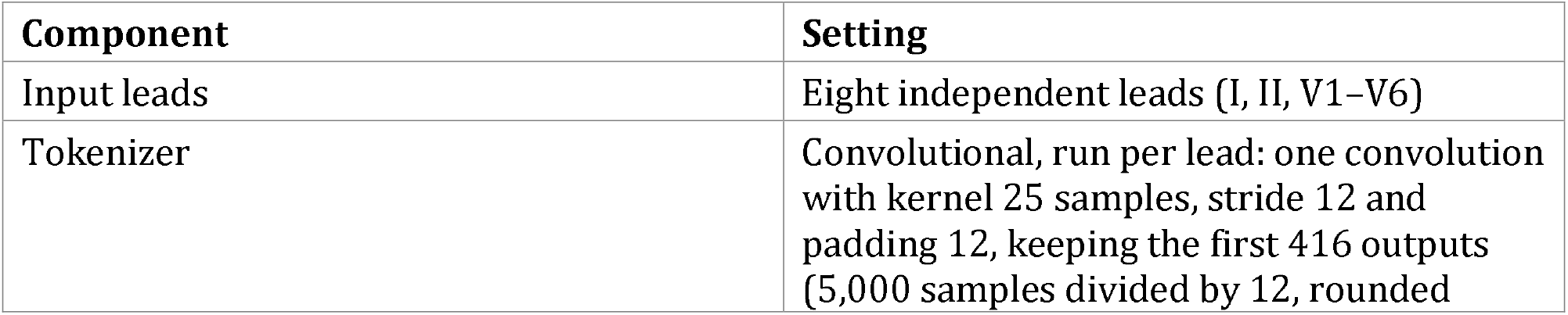
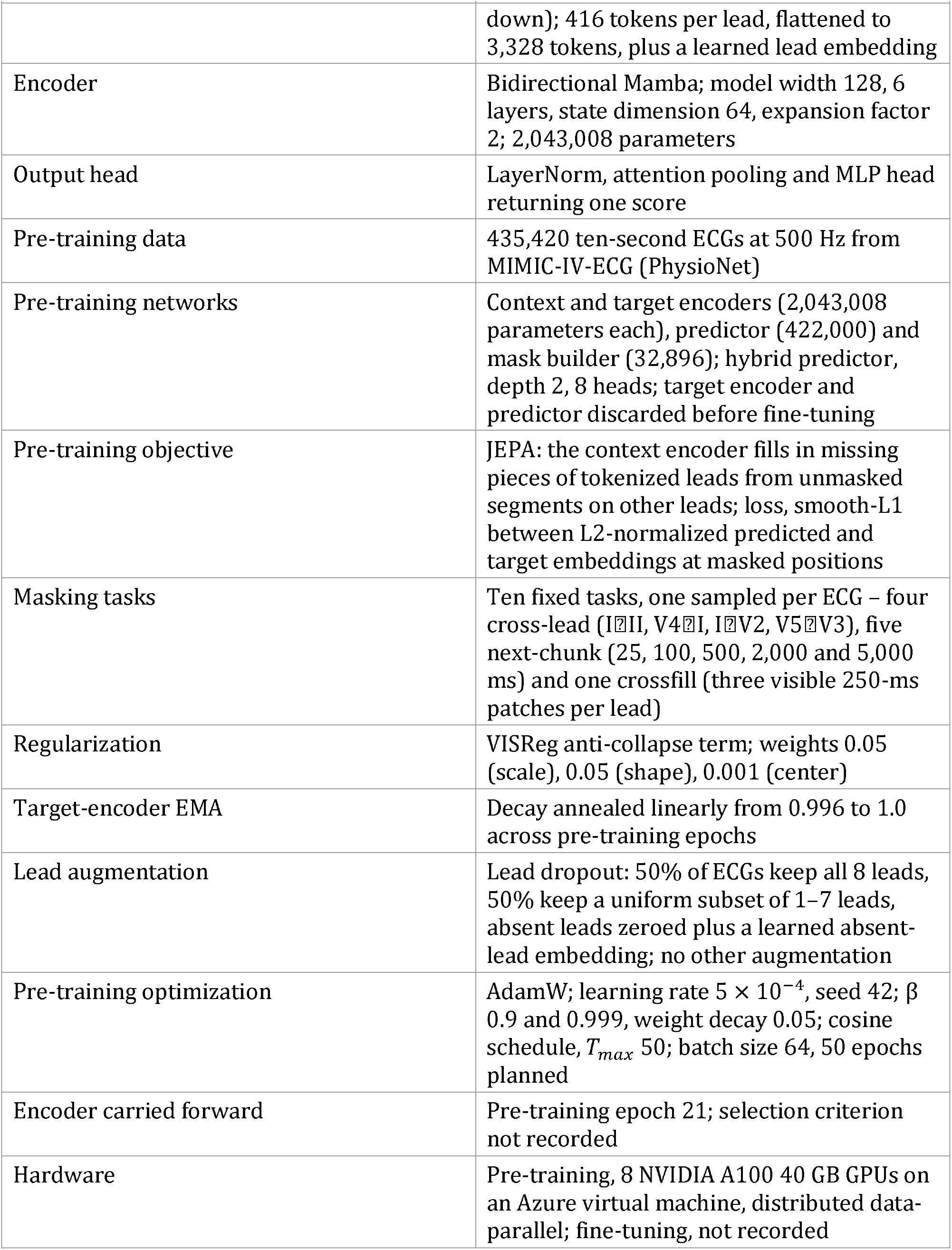
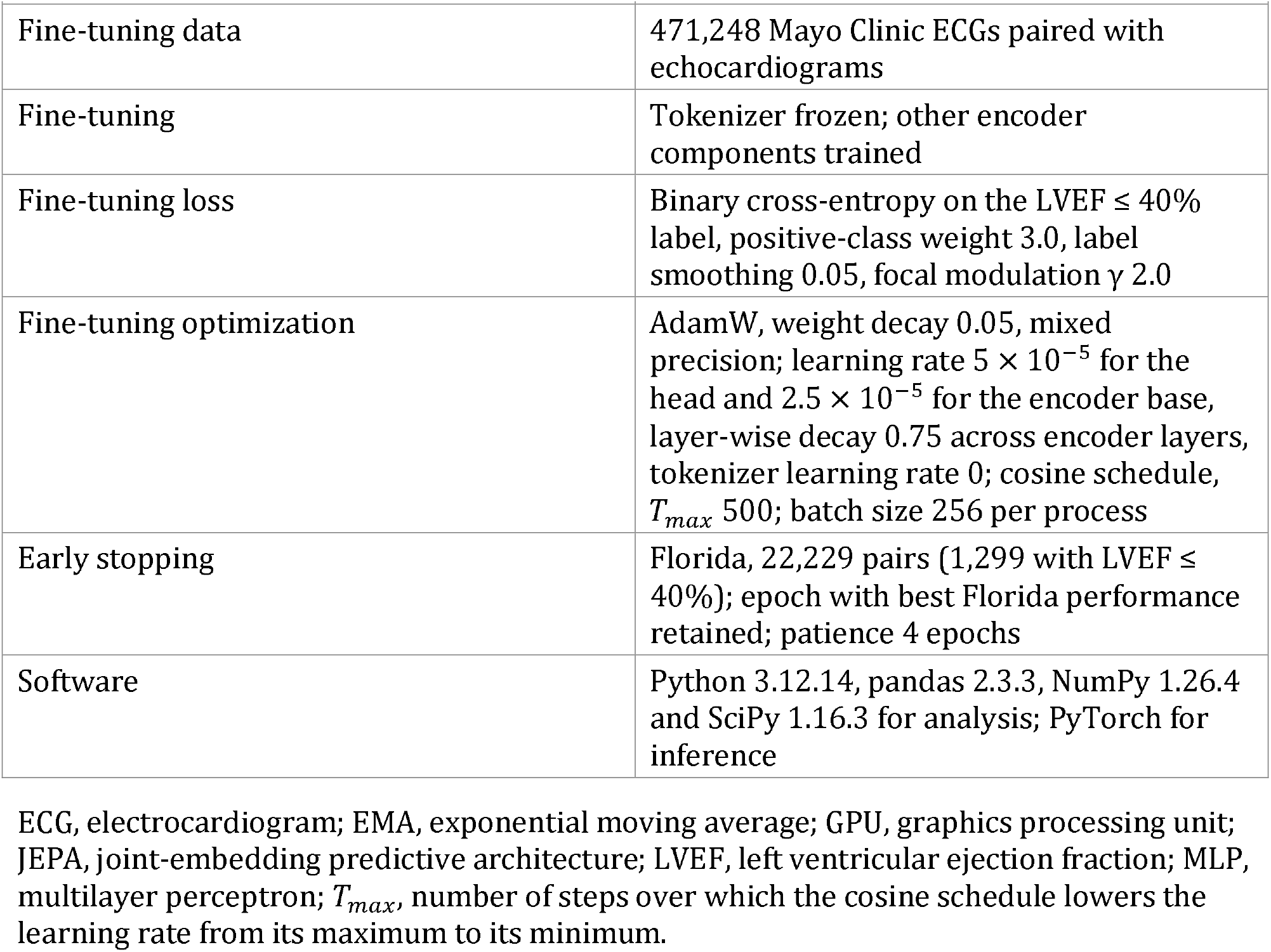
Model and training settings.

| Component | Setting |
|---|---|
| Input leads | Eight independent leads (I, II, V1–V6) |
| Tokenizer | Convolutional, run per lead: one convolution with kernel 25 samples, stride 12 and padding 12, keeping the first 416 outputs (5,000 samples divided by 12, rounded down); 416 tokens per lead, flattened to 3,328 tokens, plus a learned lead embedding |
| Encoder | Bidirectional Mamba; model width 128, 6 layers, state dimension 64, expansion factor 2; 2,043,008 parameters |
| Output head | LayerNorm, attention pooling and MLP head returning one score |
| Pre-training data | 435,420 ten-second ECGs at 500 Hz from MIMIC-IV-ECG (PhysioNet) |
| Pre-training networks | Context and target encoders (2,043,008 parameters each), predictor (422,000) and mask builder (32,896); hybrid predictor, depth 2, 8 heads; target encoder and predictor discarded before fine-tuning |
| Pre-training objective | JEPA: the context encoder fills in missing pieces of tokenized leads from unmasked segments on other leads; loss, smooth-L1 between L2-normalized predicted and target embeddings at masked positions |
| Masking tasks | Ten fixed tasks, one sampled per ECG – four cross-lead (I⍰II, V4⍰I, I⍰V2, V5⍰V3), five next-chunk (25, 100, 500, 2,000 and 5,000 ms) and one crossfill (three visible 250-ms patches per lead) |
| Regularization | VISReg anti-collapse term; weights 0.05 (scale), 0.05 (shape), 0.001 (center) |
| Target-encoder EMA | Decay annealed linearly from 0.996 to 1.0 across pre-training epochs |
| Lead augmentation | Lead dropout: 50% of ECGs keep all 8 leads, 50% keep a uniform subset of 1–7 leads, absent leads zeroed plus a learned absent-lead embedding; no other augmentation |
| Pre-training optimization | AdamW; learning rate 5 × 10<sup>−4</sup>, seed 42; β 0.9 and 0.999, weight decay 0.05; cosine schedule, *T<sub>max</sub>* 50; batch size 64, 50 epochs planned |
| Encoder carried forward | Pre-training epoch 21; selection criterion not recorded |
| Hardware | Pre-training, 8 NVIDIA A100 40 GB GPUs on an Azure virtual machine, distributed data-parallel; fine-tuning, not recorded |
| Fine-tuning data | 471,248 Mayo Clinic ECGs paired with echocardiograms |
| Fine-tuning | Tokenizer frozen; other encoder components trained |
| Fine-tuning loss | Binary cross-entropy on the LVEF ≤ 40% label, positive-class weight 3.0, label smoothing 0.05, focal modulation γ 2.0 |
| Fine-tuning optimization | AdamW, weight decay 0.05, mixed precision; learning rate 5 × 10<sup>−5</sup> for the head and 2.5 × 10<sup>−5</sup> for the encoder base, layer-wise decay 0.75 across encoder layers, tokenizer learning rate 0; cosine schedule, *T<sub>max</sub>* 500; batch size 256 per process |
| Early stopping | Florida, 22,229 pairs (1,299 with LVEF ≤ 40%); epoch with best Florida performance retained; patience 4 epochs |
| Software | Python 3.12.14, pandas 2.3.3, NumPy 1.26.4 and SciPy 1.16.3 for analysis; PyTorch for inference |

ECG, electrocardiogram; EMA, exponential moving average; GPU, graphics processing unit; JEPA, joint-embedding predictive architecture; LVEF, left ventricular ejection fraction; MLP, multilayer perceptron; *T<sub>max</sub>*, number of steps over which the cosine schedule lowers the learning rate from its maximum to its minimum.

**Supplementary Table S3.** Threshold derivation and performance targets.

| Item | Record |  |
|---|---|---|
| **A. Threshold** |  |  |
| Development units | Seven: Rochester, the five MCHS sites and Florida |  |
| Criterion per unit | Optimal F1 threshold at Rochester and Florida; optimal Youden’s J threshold at each MCHS site |  |
| Combination | Equal weight per unit, averaged to 0.350097 |  |
| Use | Fixed on Mayo development data; applied unchanged to Arizona and all external sites |  |
| Protocol wording | Youden’s J, computed once on training data; the threshold was derived by the per-unit criteria in A |  |
| Not recorded | Candidate grid; tie handling; the seven unit thresholds; lock date before the Arizona evaluation |  |
| **B. Performance targets** |  |  |
| Protocol table | Sensitivity ≥80% and specificity ≥80%, each a point estimate with two-sided 95% CI; PPV ≥25% and NPV ≥95% at cohort prevalence |  |
| Analysis-plan wording | Lower 95% confidence bounds of sensitivity and specificity at or above the targets |  |
| **Item** | **Design protocol, 07/29/2026** | **Final analysis** |
| **C. Protocol to final** |  |  |
| Pairs per patient | Main text: more than one allowed; external appendix: exactly one, smallest ECG–echo interval | External: one, closest in time; Mayo: more than one |
| Enrichment | Main text: enriched for patients at elevated risk; external appendix: characteristics not to be enriched manually | External: random sample per hospital, no enrichment |
| Prior LVEF ≤ 40% | No established LVEF ≤ 40% at the index ECG | No such exclusion in the external set |
| Directional pairing | Main text: ECG after an echocardiogram paired only if LVEF >40%; external appendix: ECG up to 15 days before or after the echocardiogram | External: any ECG within 15 days before or after; Florida and Arizona one-pair sets: the same rule, which, unlike the development pairing, admits ECGs recorded after an echocardiogram with LVEF ≤ 40% |
| Indeterminate outputs | Not scored as positive or negative in the main-text analysis | External: 156 (150 signal quality, 6 missing) excluded from the primary estimate; best- and worst-case analyses |
| Pacing | Paced-rhythm ECGs excluded | External: patients with a paced rhythm or pacemaker excluded |
| Planned analyses | Subgroups by site, sex, race/ethnicity, age group, comorbidity strata and device; calibration; LVEF method in sensitivity analyses | Post hoc: ECG–echo interval (≤ 1 and ≤ 7 days), ECG before or after echo, and 449-pair exclusion analyses |

CI, confidence interval; MCHS, Mayo Clinic Health System; NPV, PPV, negative and positive predictive value.

**Supplementary Table S4.** External cohort accounting and data checks.

| **Provider cohort construction, sites combined** | **Patients** |
|---|---|
| Adults (≥18 years) with ≥1 extracted ECG | 2,062,521 |
| ECG technical specification met | 2,002,688 |
| ECG quality screening passed | 1,975,742 |
| Non-paced ECG | 1,950,571 |
| ≥1 transthoracic echocardiogram with quantitative Simpson’s biplane LVEF | 183,469 |
| Temporal pairing rule met | 123,127 |
| Random sample, 3,500 per hospital | 17,500 |
| **Data check** | **Finding** |
| ECG–echo timing | Of 17,500 ECGs, 12,814 preceded or coincided with the echocardiogram and 4,686 followed it; 449 of those followed an echocardiogram with LVEF ≤ 40%, and all were retained |
| Indeterminate outputs | 156: 150 signal-quality failures, 6 missing outputs; 8 with LVEF ≤ 40%, 148 with LVEF >40%; 42, 13, 17, 37 and 47 at Sites 1–5 |
| Pacing | Two retained ECG interpretations suggested a possible or prior pacemaker; no device status or procedure code recorded |
| Acquisition filters | High-pass 0.05–0.56 Hz; low-pass 100 or 150 Hz |

Steps before sampling were run by the data provider for all sites combined. ECG, electrocardiogram; LVEF, left ventricular ejection fraction.

**Supplementary Table S5.** Further cohort characteristics (A), external comorbidities by reference standard (B) and ECG acquisition systems (C)

| **A. Characteristic** | **Rochester** | **MCHS** | **Florida** | **Arizona** | **External** |
|---|---|---|---|---|---|
| Index ECG dates | NA | NA | NA | NA | June 2016–April 2026 |
| LVEF, %, mean (SD) | NA | NA | NA | NA | 58.1 (12.1) |
| LVEF ≤ 30%, n (%) | NA | NA | NA | NA | 765 (4.4%) |
| LVEF 31–35%, n (%) | NA | NA | NA | NA | 382 (2.2%) |
| LVEF 36–40%, n (%) | NA | NA | NA | NA | 486 (2.8%) |
| LVEF 41–49%, n (%) | NA | NA | NA | NA | 1,290 (7.4%) |
| LVEF ≥50%, n (%) | NA | NA | NA | NA | 14,577 (83.3%) |
| ECG–echo interval, days, median; % within 1 day | NA | NA | NA | NA | 0.14; 78% |
| Age, years, median (IQR) | 63 (51–72) | 68 (58–76) | 66 (55–74) | 67 (56–75) | 68 (56–78) |
| Sex not recorded, n | 15 | 0 | 0 | 7 | 1 |
| Asian, n (%) | 7,498 (1.9%) | 651 (0.8%) | 556 (2.5%) | 988 (3.4%) | 633 (3.6%) |
| Native American or Pacific Islander, n (%) | 2,554 (0.7%) | 330 (0.4%) | 91 (0.4%) | 686 (2.3%) | — |
| American Indian or Alaska Native, n (%) | — | — | — | — | 33 (0.2%) |
| Native Hawaiian or Pacific Islander, n (%) | — | — | — | — | 59 (0.3%) |
| Other race, n (%) | 4,838 (1.3%) | 347 (0.4%) | 174 (0.8%) | 344 (1.2%) | 0 (0.0%) |
| Race not recorded, n (%) | 6,163 (1.6%) | 469 (0.6%) | 377 (1.7%) | 346 (1.2%) | 1,899 (10.9%) |
| Not Hispanic or Latino, n (%) | 363,201 (93.9%) | 81,816 (97.1%) | 20,636 (92.8%) | 26,193 (89.4%) | 15,488 (88.5%) |
| Ethnicity not recorded, n (%) | 15,312 (4.0%) | 937 (1.1%) | 521 (2.3%) | 674 (2.3%) | 402 (2.3%) |
| Prior myocardial infarction, n (%) | 36,814 (9.5%) | 13,890 (16.5%) | 1,934 (8.7%) | 3,425 (11.7%) | 2,420 (13.8%) |
| Cardiomyopathy, n (%) | 39,830 (10.3%) | 7,022 (8.3%) | 2,163 (9.7%) | 3,223 (11.0%) | 1,553 (8.9%) |
| Diabetes mellitus, n (%) | 52,369 (13.5%) | 20,468 (24.3%) | 4,473 (20.1%) | 5,689 (19.4%) | 5,813 (33.2%) |
| Hypertension, n (%) | 138,552 (35.8%) | 44,810 (53.2%) | 11,890 (53.5%) | 15,256 (52.1%) | 13,025 (74.4%) |
| AF, persistent or chronic, n (%) | 74,457 (19.2%) | 18,290 (21.7%) | 4,651 (20.9%) | 5,798 (19.8%) | 1,324 (7.6%) |
| AF, paroxysmal or unspecified, n (%) | 45,938 (11.9%) | 10,941 (13.0%) | 1,583 (7.1%) | 2,242 (7.7%) | — |
| AF, paroxysmal, n (%) | — | — | — | — | 1,545 (8.8%) |
| AF, unspecified, n (%) | — | — | — | — | 1,243 (7.1%) |
| CKD stage 3–5, n (%) | 35,099 (9.1%) | 12,622 (15.0%) | 3,956 (17.8%) | 5,115 (17.5%) | 2,978 (17.0%) |
| End-stage renal disease, n (%) | NA | NA | NA | NA | 741 (4.2%) |
| Coronary artery disease, n (%) | 103,222 (26.7%) | 25,807 (30.6%) | 7,143 (32.1%) | 9,025 (30.8%) | 6,399 (36.6%) |
| Heart failure diagnosis, n (%) | NA | NA | NA | NA | 5,046 (28.8%) |
| **B. External cohort** | **LVEF ≤ 40%** | **LVEF >40%** |  |  |  |
| **LVEF band, n (%) of delivered pairs with LVEF ≤ 40% (n = 1,633)** |  |  |  |  |  |
| ≤ 30% | 765 (46.8%) | — |  |  |  |
| 31–35% | 382 (23.4%) | — |  |  |  |
| 36–40% | 486 (29.8%) | — |  |  |  |
| **Documented comorbidity, % of analyzed pairs (n = 1,625 and 15,719)** |  |  |  |  |  |
| AF, persistent or chronic | 10.3% | 7.3% |  |  |  |
| AF, paroxysmal | 10.0% | 8.7% |  |  |  |
| AF, unspecified | 11.4% | 6.7% |  |  |  |
| Cardiomyopathy | 36.6% | 6.1% |  |  |  |
| End-stage renal disease | 6.6% | 4.0% |  |  |  |
| Hypertension | 77.4% | 74.2% |  |  |  |
| Diabetes mellitus | 42.2% | 32.4% |  |  |  |
| Coronary artery disease | 50.5% | 35.3% |  |  |  |
| Heart failure diagnosis | 67.2% | 25.0% |  |  |  |
| **C. ECG acquisition system, n (%)** | **Mayo cohorts combined** | **External** |  |  |  |
| GE MAC 5500/5500 HD | 317,177 (60.7%) | 7,961 (45.5%) |  |  |  |
| GE MAC VU360 | 182,912 (35.0%) | 3,184 (18.2%) |  |  |  |
| GE MAC 5000 | 14,571 (2.8%) | — |  |  |  |
| GE MAC 800 | 2,290 (0.4%) | — |  |  |  |
| Other GE MAC | 3,900 (0.7%) | — |  |  |  |
| GE MAC 5 | — | 34 (0.2%) |  |  |  |
| Philips PageWriter TC70 | — | 6,321 (36.1%) |  |  |  |
| Other or unknown | 1,916 (0.4%) | — |  |  |  |

Mayo percentages are of pairs; external percentages are of patients, one pair each. Comorbidity codes are defined in Supplementary Table S1. —, not reported as a separate category. AF, atrial fibrillation; CKD, chronic kidney disease; ECG, electrocardiogram; echo, echocardiogram; IQR, interquartile range; LVEF, left ventricular ejection fraction; MCHS, Mayo Clinic Health System; NA, not available in the aggregate Mayo cohort data; SD, standard deviation.

**Supplementary Table S6.**
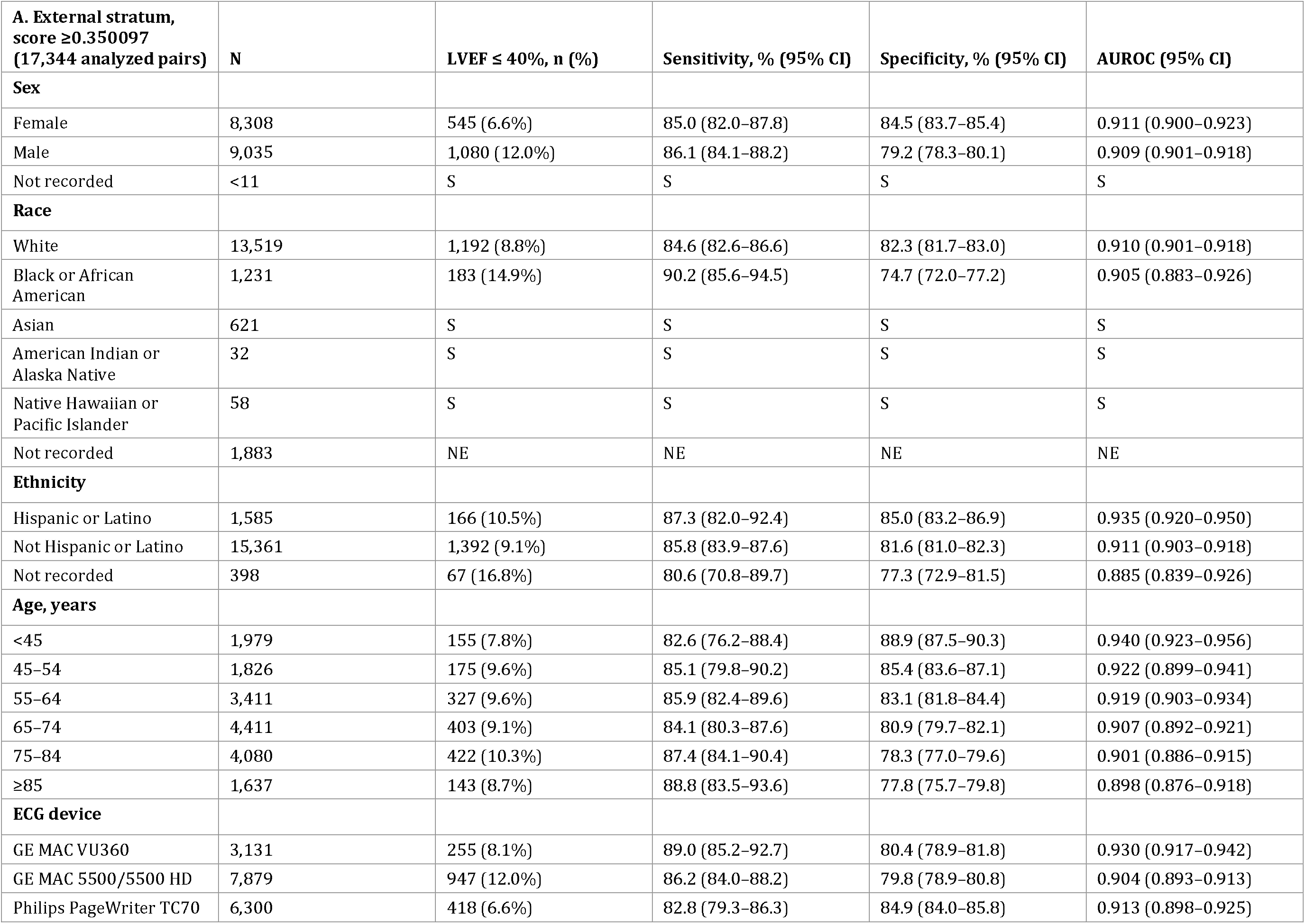
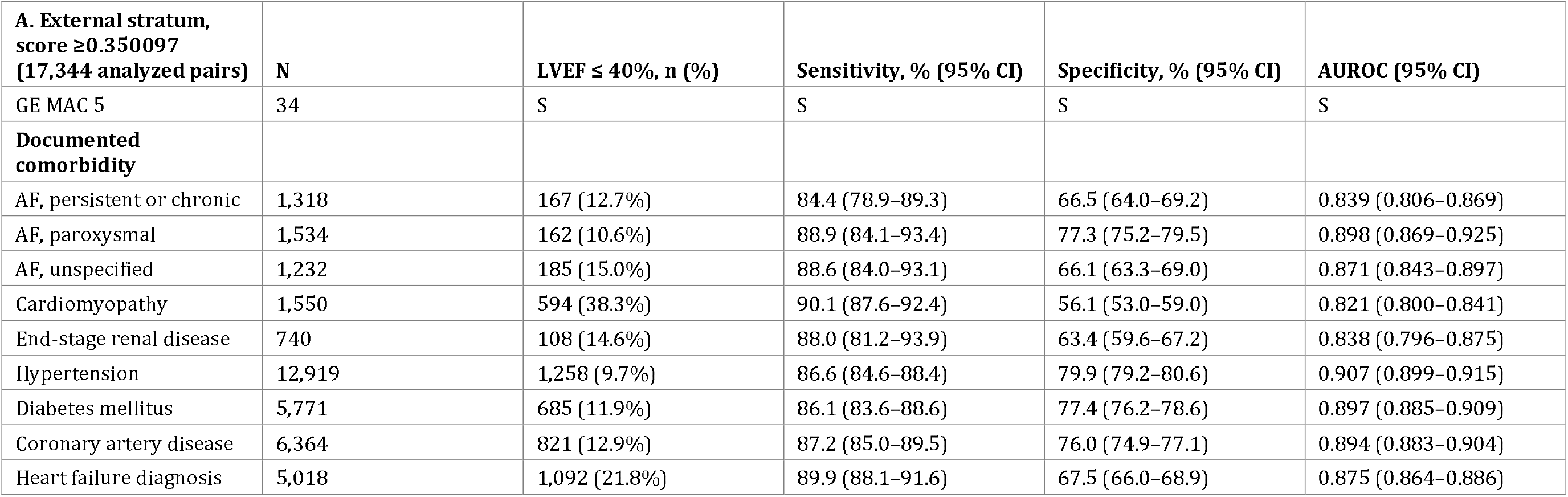
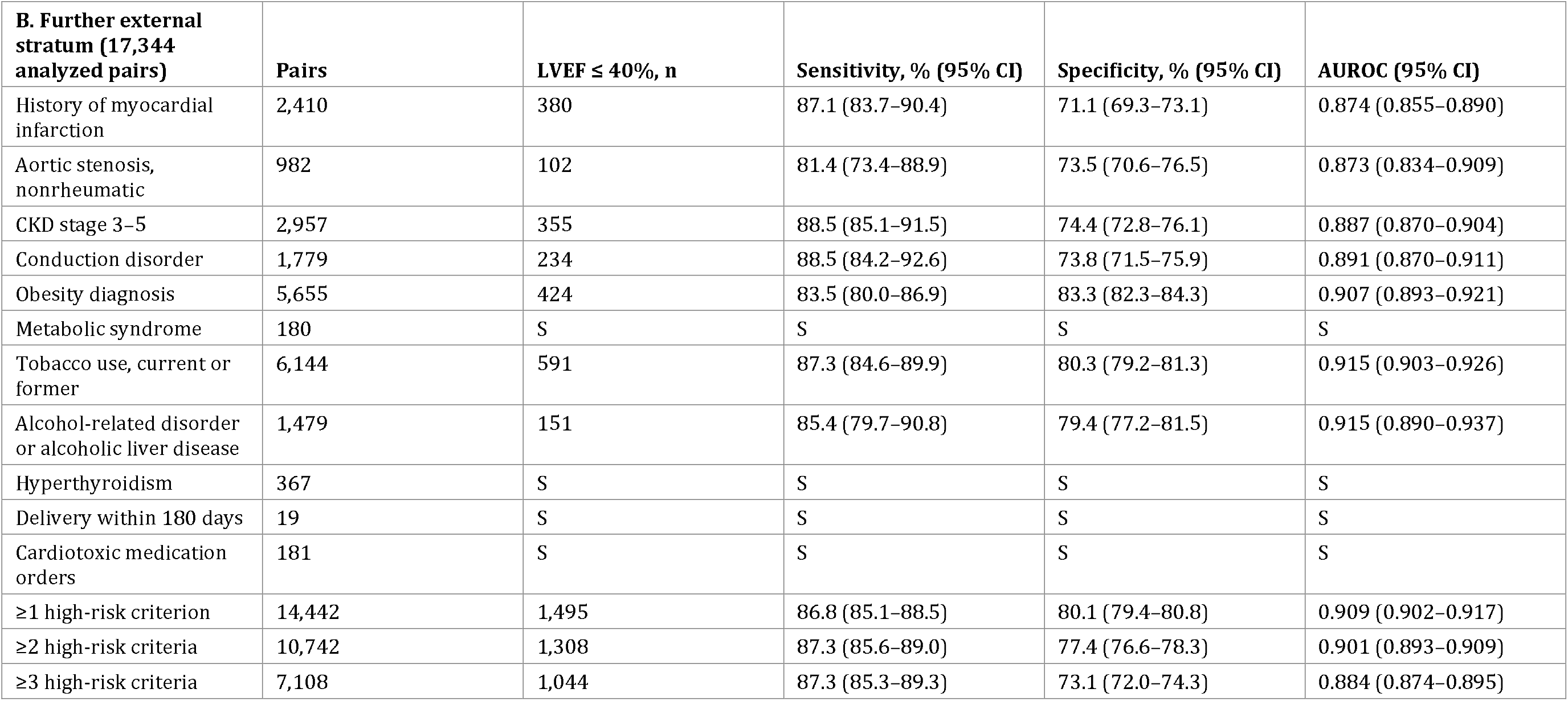
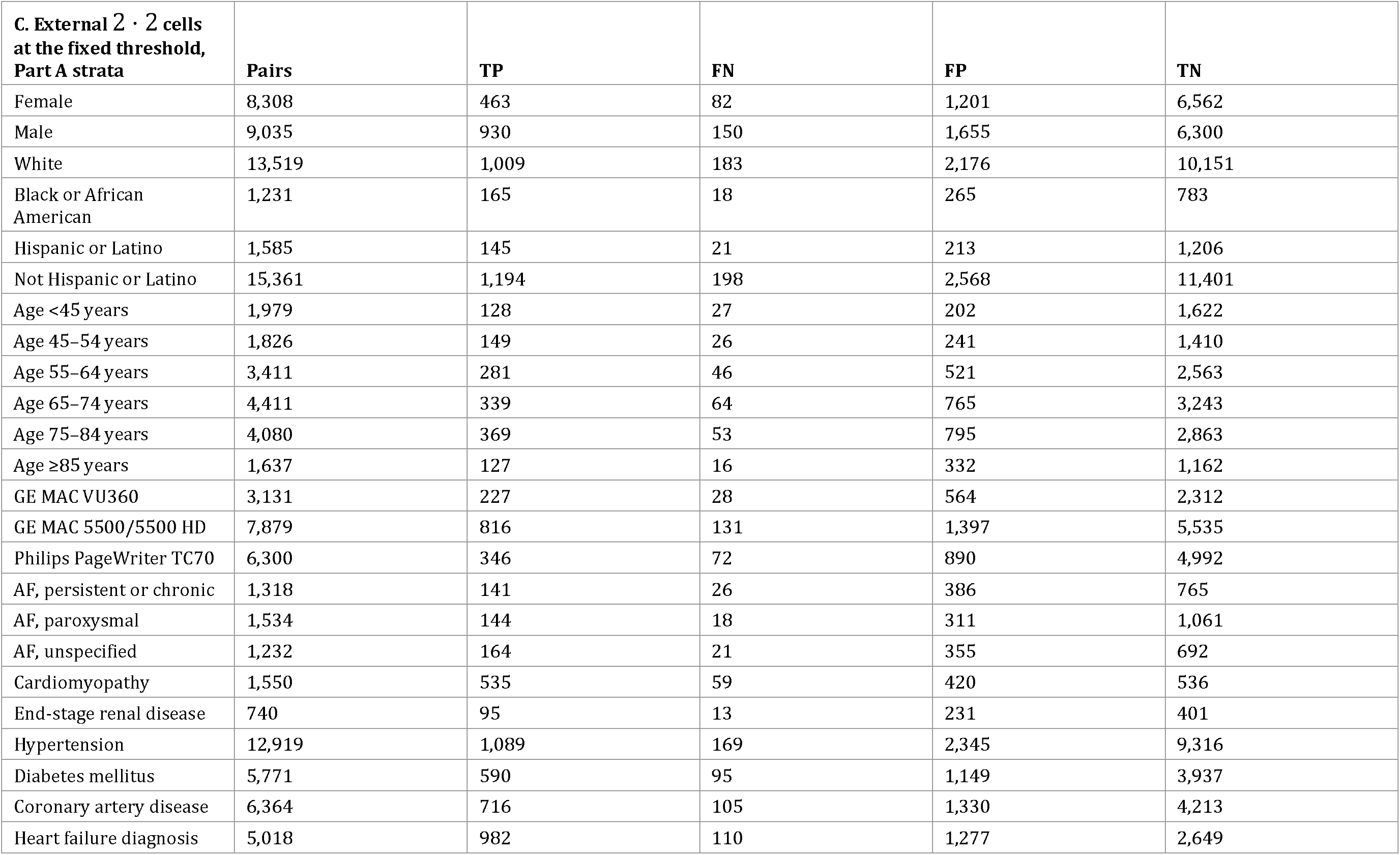
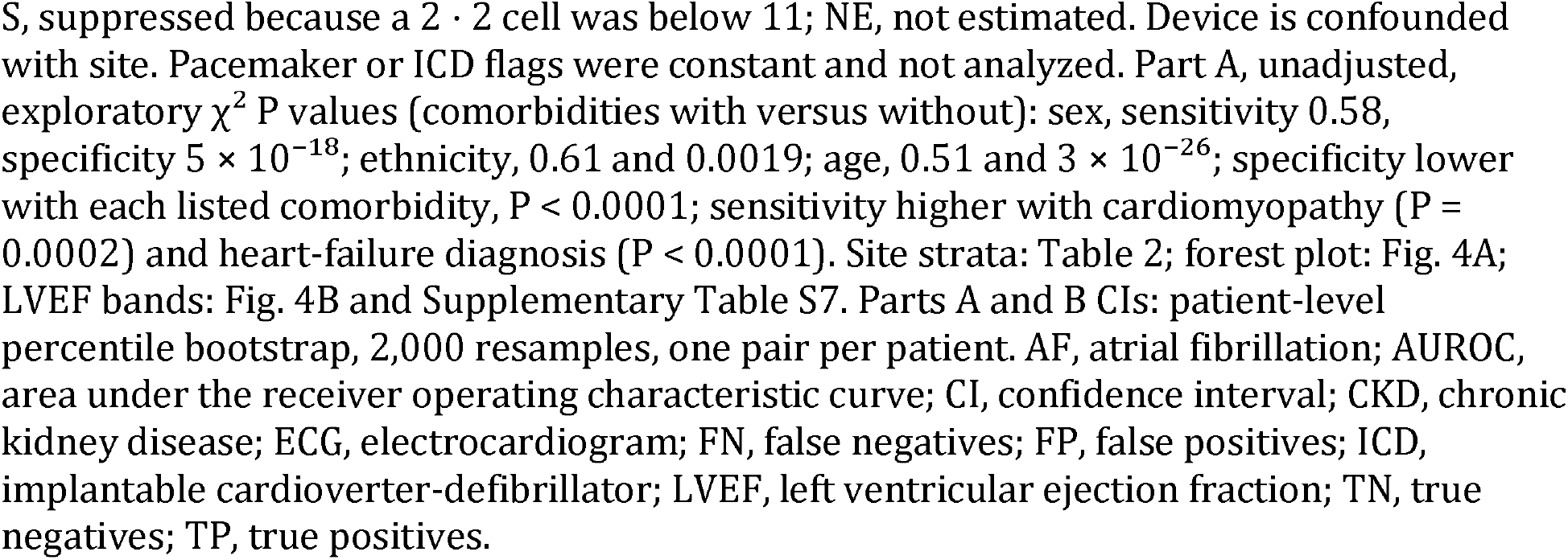
Subgroup results: external strata (A, B) and external 2 2 cells (C)

| **A. External stratum, score ≥0.350097 (17,344 analyzed pairs)** | **N** | **LVEF ≤ 40%, n (%)** | **Sensitivity, % (95% CI)** | **Specificity, % (95% CI)** | **AUROC (95% CI)** |
|---|---|---|---|---|---|
| **Sex** |  |  |  |  |  |
| Female | 8,308 | 545 (6.6%) | 85.0 (82.0–87.8) | 84.5 (83.7–85.4) | 0.911 (0.900–0.923) |
| Male | 9,035 | 1,080 (12.0%) | 86.1 (84.1–88.2) | 79.2 (78.3–80.1) | 0.909 (0.901–0.918) |
| Not recorded | <11 | S | S | S | S |
| **Race** |  |  |  |  |  |
| White | 13,519 | 1,192 (8.8%) | 84.6 (82.6–86.6) | 82.3 (81.7–83.0) | 0.910 (0.901–0.918) |
| Black or African American | 1,231 | 183 (14.9%) | 90.2 (85.6–94.5) | 74.7 (72.0–77.2) | 0.905 (0.883–0.926) |
| Asian | 621 | S | S | S | S |
| American Indian or Alaska Native | 32 | S | S | S | S |
| Native Hawaiian or Pacific Islander | 58 | S | S | S | S |
| Not recorded | 1,883 | NE | NE | NE | NE |
| **Ethnicity** |  |  |  |  |  |
| Hispanic or Latino | 1,585 | 166 (10.5%) | 87.3 (82.0–92.4) | 85.0 (83.2–86.9) | 0.935 (0.920–0.950) |
| Not Hispanic or Latino | 15,361 | 1,392 (9.1%) | 85.8 (83.9–87.6) | 81.6 (81.0–82.3) | 0.911 (0.903–0.918) |
| Not recorded | 398 | 67 (16.8%) | 80.6 (70.8–89.7) | 77.3 (72.9–81.5) | 0.885 (0.839–0.926) |
| **Age, years** |  |  |  |  |  |
| <45 | 1,979 | 155 (7.8%) | 82.6 (76.2–88.4) | 88.9 (87.5–90.3) | 0.940 (0.923–0.956) |
| 45–54 | 1,826 | 175 (9.6%) | 85.1 (79.8–90.2) | 85.4 (83.6–87.1) | 0.922 (0.899–0.941) |
| 55–64 | 3,411 | 327 (9.6%) | 85.9 (82.4–89.6) | 83.1 (81.8–84.4) | 0.919 (0.903–0.934) |
| 65–74 | 4,411 | 403 (9.1%) | 84.1 (80.3–87.6) | 80.9 (79.7–82.1) | 0.907 (0.892–0.921) |
| 75–84 | 4,080 | 422 (10.3%) | 87.4 (84.1–90.4) | 78.3 (77.0–79.6) | 0.901 (0.886–0.915) |
| ≥85 | 1,637 | 143 (8.7%) | 88.8 (83.5–93.6) | 77.8 (75.7–79.8) | 0.898 (0.876–0.918) |
| **ECG device** |  |  |  |  |  |
| GE MAC VU360 | 3,131 | 255 (8.1%) | 89.0 (85.2–92.7) | 80.4 (78.9–81.8) | 0.930 (0.917–0.942) |
| GE MAC 5500/5500 HD | 7,879 | 947 (12.0%) | 86.2 (84.0–88.2) | 79.8 (78.9–80.8) | 0.904 (0.893–0.913) |
| Philips PageWriter TC70 | 6,300 | 418 (6.6%) | 82.8 (79.3–86.3) | 84.9 (84.0–85.8) | 0.913 (0.898–0.925) |
| **A. External stratum, score ≥0.350097 (17,344 analyzed pairs)** | **N** | **LVEF ≤ 40%, n (%)** | **Sensitivity, % (95% CI)** | **Specificity, % (95% CI)** | **AUROC (95% CI)** |
| GE MAC 5 | 34 | S | S | S | S |
| **Documented comorbidity** |  |  |  |  |  |
| AF, persistent or chronic | 1,318 | 167 (12.7%) | 84.4 (78.9–89.3) | 66.5 (64.0–69.2) | 0.839 (0.806–0.869) |
| AF, paroxysmal | 1,534 | 162 (10.6%) | 88.9 (84.1–93.4) | 77.3 (75.2–79.5) | 0.898 (0.869–0.925) |
| AF, unspecified | 1,232 | 185 (15.0%) | 88.6 (84.0–93.1) | 66.1 (63.3–69.0) | 0.871 (0.843–0.897) |
| Cardiomyopathy | 1,550 | 594 (38.3%) | 90.1 (87.6–92.4) | 56.1 (53.0–59.0) | 0.821 (0.800–0.841) |
| End-stage renal disease | 740 | 108 (14.6%) | 88.0 (81.2–93.9) | 63.4 (59.6–67.2) | 0.838 (0.796–0.875) |
| Hypertension | 12,919 | 1,258 (9.7%) | 86.6 (84.6–88.4) | 79.9 (79.2–80.6) | 0.907 (0.899–0.915) |
| Diabetes mellitus | 5,771 | 685 (11.9%) | 86.1 (83.6–88.6) | 77.4 (76.2–78.6) | 0.897 (0.885–0.909) |
| Coronary artery disease | 6,364 | 821 (12.9%) | 87.2 (85.0–89.5) | 76.0 (74.9–77.1) | 0.894 (0.883–0.904) |
| Heart failure diagnosis | 5,018 | 1,092 (21.8%) | 89.9 (88.1–91.6) | 67.5 (66.0–68.9) | 0.875 (0.864–0.886) |
| **B. Further external stratum (17,344 analyzed pairs)** | **Pairs** | **LVEF ≤ 40%, n** | **Sensitivity, % (95% CI)** | **Specificity, % (95% CI)** | **AUROC (95% CI)** |
| History of myocardial infarction | 2,410 | 380 | 87.1 (83.7–90.4) | 71.1 (69.3–73.1) | 0.874 (0.855–0.890) |
| Aortic stenosis, nonrheumatic | 982 | 102 | 81.4 (73.4–88.9) | 73.5 (70.6–76.5) | 0.873 (0.834–0.909) |
| CKD stage 3–5 | 2,957 | 355 | 88.5 (85.1–91.5) | 74.4 (72.8–76.1) | 0.887 (0.870–0.904) |
| Conduction disorder | 1,779 | 234 | 88.5 (84.2–92.6) | 73.8 (71.5–75.9) | 0.891 (0.870–0.911) |
| Obesity diagnosis | 5,655 | 424 | 83.5 (80.0–86.9) | 83.3 (82.3–84.3) | 0.907 (0.893–0.921) |
| Metabolic syndrome | 180 | S | S | S | S |
| Tobacco use, current or former | 6,144 | 591 | 87.3 (84.6–89.9) | 80.3 (79.2–81.3) | 0.915 (0.903–0.926) |
| Alcohol-related disorder or alcoholic liver disease | 1,479 | 151 | 85.4 (79.7–90.8) | 79.4 (77.2–81.5) | 0.915 (0.890–0.937) |
| Hyperthyroidism | 367 | S | S | S | S |
| Delivery within 180 days | 19 | S | S | S | S |
| Cardiotoxic medication orders | 181 | S | S | S | S |
| ≥1 high-risk criterion | 14,442 | 1,495 | 86.8 (85.1–88.5) | 80.1 (79.4–80.8) | 0.909 (0.902–0.917) |
| ≥2 high-risk criteria | 10,742 | 1,308 | 87.3 (85.6–89.0) | 77.4 (76.6–78.3) | 0.901 (0.893–0.909) |
| ≥3 high-risk criteria | 7,108 | 1,044 | 87.3 (85.3–89.3) | 73.1 (72.0–74.3) | 0.884 (0.874–0.895) |
| **C. External 2 · 2 cells at the fixed threshold, Part A strata** | **Pairs** | **TP** | **FN** | **FP** | **TN** |
| Female | 8,308 | 463 | 82 | 1,201 | 6,562 |
| Male | 9,035 | 930 | 150 | 1,655 | 6,300 |
| White | 13,519 | 1,009 | 183 | 2,176 | 10,151 |
| Black or African American | 1,231 | 165 | 18 | 265 | 783 |
| Hispanic or Latino | 1,585 | 145 | 21 | 213 | 1,206 |
| Not Hispanic or Latino | 15,361 | 1,194 | 198 | 2,568 | 11,401 |
| Age <45 years | 1,979 | 128 | 27 | 202 | 1,622 |
| Age 45–54 years | 1,826 | 149 | 26 | 241 | 1,410 |
| Age 55–64 years | 3,411 | 281 | 46 | 521 | 2,563 |
| Age 65–74 years | 4,411 | 339 | 64 | 765 | 3,243 |
| Age 75–84 years | 4,080 | 369 | 53 | 795 | 2,863 |
| Age ≥85 years | 1,637 | 127 | 16 | 332 | 1,162 |
| GE MAC VU360 | 3,131 | 227 | 28 | 564 | 2,312 |
| GE MAC 5500/5500 HD | 7,879 | 816 | 131 | 1,397 | 5,535 |
| Philips PageWriter TC70 | 6,300 | 346 | 72 | 890 | 4,992 |
| AF, persistent or chronic | 1,318 | 141 | 26 | 386 | 765 |
| AF, paroxysmal | 1,534 | 144 | 18 | 311 | 1,061 |
| AF, unspecified | 1,232 | 164 | 21 | 355 | 692 |
| Cardiomyopathy | 1,550 | 535 | 59 | 420 | 536 |
| End-stage renal disease | 740 | 95 | 13 | 231 | 401 |
| Hypertension | 12,919 | 1,089 | 169 | 2,345 | 9,316 |
| Diabetes mellitus | 5,771 | 590 | 95 | 1,149 | 3,937 |
| Coronary artery disease | 6,364 | 716 | 105 | 1,330 | 4,213 |
| Heart failure diagnosis | 5,018 | 982 | 110 | 1,277 | 2,649 |

S, suppressed because a 2 · 2 cell was below 11; NE, not estimated. Device is confounded with site. Pacemaker or ICD flags were constant and not analyzed. Part A, unadjusted, exploratory χ<sup>2</sup> P values (comorbidities with versus without): sex, sensitivity 0.58, specificity 5 × 10<sup>−18</sup>; ethnicity, 0.61 and 0.0019; age, 0.51 and 3 × 10<sup>−26</sup>; specificity lower with each listed comorbidity, P < 0.0001; sensitivity higher with cardiomyopathy (P = 0.0002) and heart-failure diagnosis (P < 0.0001). Site strata: Table 2; forest plot: Fig. 4A; LVEF bands: Fig. 4B and Supplementary Table S7. Parts A and B CIs: patient-level percentile bootstrap, 2,000 resamples, one pair per patient. AF, atrial fibrillation; AUROC, area under the receiver operating characteristic curve; CI, confidence interval; CKD, chronic kidney disease; ECG, electrocardiogram; FN, false negatives; FP, false positives; ICD, implantable cardioverter-defibrillator; LVEF, left ventricular ejection fraction; TN, true negatives; TP, true positives.

**Supplementary Table S7.** LVEF bands and calibration, external cohort.

| LVEF band, % | Analysed pairs | Test-positive | Test-negative | Indeterminate | Test-positive, % (95% CI) |
|---|---|---|---|---|---|
| ≤ 20 | 240 | 229 | 11 | <11 | 95.4 (91.9–97.7) |
| >20 to ≤ 25 | 216 | 196 | 20 | <11 | 90.7 (86.1–94.3) |
| >25 to ≤ 30 | 305 | 277 | 28 | <11 | 90.8 (87.0–93.8) |
| >30 to ≤ 35 | 381 | 311 | 70 | <11 | 81.6 (77.4–85.4) |
| >35 to ≤ 40 | 483 | 380 | 103 | <11 | 78.7 (74.7–82.2) |
| >40 to ≤ 45 | 615 | 417 | 198 | <11 | 67.8 (64.0–71.5) |
| >45 to ≤ 50 | 911 | 475 | 436 | <11 | 52.1 (48.8–55.4) |
| >50 to ≤ 55 | 1,909 | 549 | 1,360 | 20 | 28.8 (26.7–30.8) |
| >55 to ≤ 60 | 3,480 | 554 | 2,926 | 29 | 15.9 (14.7–17.2) |
| >60 to ≤ 65 | 4,214 | 447 | 3,767 | 38 | 10.6 (9.7–11.6) |
| >65 | 4,590 | 414 | 4,176 | 49 | 9.0 (8.2–9.9) |
| **Score decile** | **Pairs** | **Mean score** | **Observed prevalence of LVEF ≤ 40%** |  |  |
| 1–3, merged | 5,203 | 0.0655 | 0.0033 |  |  |
| 4 | 1,735 | 0.1295 | 0.0069 |  |  |
| 5 | 1,734 | 0.1705 | 0.0110 |  |  |
| 6 | 1,734 | 0.2182 | 0.0265 |  |  |
| 7 | 1,735 | 0.2748 | 0.0363 |  |  |
| 8 | 1,734 | 0.3471 | 0.0963 |  |  |
| 9 | 1,734 | 0.4478 | 0.2065 |  |  |
| 10 | 1,735 | 0.6188 | 0.5435 |  |  |
| **Score summary** | **Value** |  |  |  |  |
| Mean (SD) | 0.2403 (0.1786) |  |  |  |  |
| Median (IQR) | 0.1928 (0.0951–0.3462) |  |  |  |  |
| Test-positive, n (%) | 4,249 (24.5%) |  |  |  |  |
| Observed prevalence of LVEF ≤ 40% | 0.0937 |  |  |  |  |
| Brier score | 0.084 |  |  |  |  |
| Calibration intercept | −0.900 |  |  |  |  |
| Calibration slope | 2.178 |  |  |  |  |

Calibration intercept and slope are from logistic regression on the logit score; 95% CIs were not computed for the Brier score, intercept or slope. CIs are exact Clopper–Pearson. Reference standard: LVEF ≤ 40% positive and >40% negative, each class subdivided by the bands. <11, count of 0–10, suppressed; CI, confidence interval; IQR, interquartile range; LVEF, left ventricular ejection fraction; SD, standard deviation.

**Supplementary Table S8.** External sensitivity analyses at the fixed threshold.

| **Analysis** | **LVEF ≤ 40%/pairs** | **TP/FN/FP/TN** | **Sensitivity, % (95% CI)** | **Specificity, % (95% CI)** | **PPV, % (95% CI)** | **NPV, % (95% CI)** | **AUROC (95% CI)** |
|---|---|---|---|---|---|---|---|
| ECG before or at echo only | 1,179/12,688 | 1,016/163/2,070/9,439 | 86.2 (84.2–88.1) | 82.0 (81.3–82.7) | 32.9 (31.3–34.6) | 98.3 (98.0–98.6) | 0.914 (0.906–0.922) |
| ECG–echo interval ≤ 1 day | 1,309/13,546 | 1,149/160/2,320/9,917 | 87.8 (85.9–89.4) | 81.0 (80.3–81.7) | 33.1 (31.6–34.7) | 98.4 (98.2–98.7) | 0.918 (0.910–0.925) |
| ECG–echo interval ≤ 7 days | 1,577/16,284 | 1,360/217/2,754/11,953 | 86.2 (84.6–87.9) | 81.3 (80.6–81.9) | 33.1 (31.6–34.6) | 98.2 (98.0–98.4) | 0.913 (0.906–0.920) |
| ECG after echo | 446/4,656 | 377/69/786/3,424 | 84.5 (81.0–87.9) | 81.3 (80.1–82.5) | 32.4 (29.6–35.0) | 98.0 (97.5–98.5) | 0.907 (0.893–0.920) |
| Indeterminates all positive | 1,633/17,500 | 1,401/232/3,004/12,863 | 85.8 (84.1–87.4) | 81.1 (80.5–81.7) | 31.8 (30.5–33.2) | 98.2 (98.0–98.5) | — |
| Indeterminates all negative | 1,633/17,500 | 1,393/240/2,856/13,011 | 85.3 (83.6–87.0) | 82.0 (81.4–82.6) | 32.8 (31.4–34.1) | 98.2 (98.0–98.4) | — |
| Missing scores imputed as the reference class | 1,633/17,500 | — | — | — | — | — | 0.9128 |
| Missing scores imputed as its complement | 1,633/17,500 | — | — | — | — | — | 0.9119 |
| All observed scores | —/17,494 | — | — | — | — | — | 0.9127 |
| **LVEF method stratum** | **Pairs** | **Sensitivity, % (95% CI)** | **Specificity, % (95% CI)** | **AUROC (95% CI)** |  |  |  |
| Simpson’s method of disks named | 8,655 | 85.2 (83.0–87.4) | 80.7 (79.8–81.5) | 0.904 (0.894–0.913) |  |  |  |
| Biplane, method not named | 8,689 | 86.4 (83.7–89.0) | 82.9 (82.1–83.8) | 0.923 (0.913–0.932) |  |  |  |
| **Site, indeterminates counted on its 3,500 pairs (point estimates, no CI)** | **Sensitivity, best / worst case, %** | **Specificity, best / worst case, %** |  |  |  |  |  |
| Site 1 | 90.3 / 89.8 | 83.7 / 82.4 |  |  |  |  |  |
| Site 2 | 87.3 / 87.3 | 85.6 / 85.2 |  |  |  |  |  |
| Site 3 | 79.6 / 79.2 | 82.7 / 82.2 |  |  |  |  |  |
| Site 4 | 83.6 / 83.3 | 81.1 / 80.0 |  |  |  |  |  |
| Site 5 | 87.4 / 86.5 | 76.4 / 74.9 |  |  |  |  |  |

Other sensitivity analyses are in Table 2; indeterminate outputs are counted in Fig. 1 and Supplementary Table S4. The two indeterminate rows count every indeterminate output as positive or as negative. In the last three rows of the first table, the quality-rejected records keep their observed scores. CIs: percentile bootstrap, 2,000 resamples; one pair per patient. —, not computed. AUROC, area under the receiver operating characteristic curve; CI, confidence interval; ECG, electrocardiogram; FN, FP, TN, TP, false/true negatives and positives; LVEF, left ventricular ejection fraction; NPV, PPV, negative/positive predictive value.

**Supplementary Table S9.** Matched comparison with a published external test at LVEF ≤ 40%.

| **Item** | **This study, external cohort** | **Carter et al., external test** |
|---|---|---|
| Outcome | LVEF ≤ 40% on transthoracic echocardiography | LVEF ≤ 40%[13] |
| Threshold | Fixed on Mayo development data, applied unchanged | Selected before the study, not adjusted[13] |
| Cohort | 5 hospitals; one ECG–echo pair per patient within 15 days | 4 U.S. sites; ECG and transthoracic echocardiogram within 30 days[13] |
| Starting, excluded and analyzed, n | 17,500 pairs delivered; 156 indeterminate excluded (150 signal quality, 6 missing output, waveform present); 17,344 analyzed | 16,000 subjects extracted; 13,960 evaluable[13] |
| Prevalence | 9.4% | 7.9%[13] |
| Sensitivity, % (95% CI) | 85.7 (84.1–87.4) | 84.5 (82.2–86.6)[13] |
| Specificity, % (95% CI) | 81.8 (81.3–82.4) | 83.6 (82.9–84.2)[13] |
| PPV, % (95% CI) | 32.8 (31.4–34.1) | 30.5 (28.8–32.1)[13] |
| NPV, % (95% CI) | 98.2 (98.0–98.5) | 98.4 (98.2–98.7)[13] |
| AUROC (95% CI) | 0.912 (0.906–0.919) | 0.919 (0.910–0.927)[13] |

Estimates are on the analyzed (evaluable) sets; best- and worst-case results for this study’s 156 indeterminate outputs are in Supplementary Table S8. Intervals: this study, patient-level percentile bootstrap, 2,000 resamples; Carter et al., exact 95% CIs[13]. AUROC, area under the receiver operating characteristic curve; CI, confidence interval; ECG, electrocardiogram; echo, echocardiogram; LVEF, left ventricular ejection fraction; NPV, PPV, negative and positive predictive value; U.S., United States; —, not reported.

## References

1. Bozkurt B, Ahmad T, Alexander K, Baker WL, Bosak K, Breathett K, et al. HF STATS 2024: heart failure epidemiology and outcomes statistics. An updated 2024 report from the Heart Failure Society of America. J Card Fail. 2025;31(1):66–116. doi:10.1016/j.cardfail.2024.07.001.

2. Wang TJ, Levy D, Benjamin EJ, Vasan RS. The epidemiology of “asymptomatic” left ventricular systolic dysfunction: implications for screening. Ann Intern Med. 2003;138(11):907–16. doi:10.7326/0003-4819-138-11-200306030-00012.

3. The SOLVD Investigators. Effect of enalapril on mortality and the development of heart failure in asymptomatic patients with reduced left ventricular ejection fractions. N Engl J Med. 1992;327(10):685–91. doi:10.1056/NEJM199209033271003.

4. Heidenreich PA, Bozkurt B, Aguilar D, Allen LA, Byun JJ, Colvin MM, et al. 2022 AHA/ACC/HFSA guideline for the management of heart failure: a report of the American College of Cardiology/American Heart Association Joint Committee on Clinical Practice Guidelines. Circulation. 2022;145(18):e895–e1032. doi:10.1161/CIR.0000000000001063.

5. Ledwidge M, Gallagher J, Conlon C, Tallon E, O’Connell E, Dawkins I, et al. Natriuretic peptide-based screening and collaborative care for heart failure: the STOP-HF randomized trial. JAMA. 2013;310(1):66–74. doi:10.1001/jama.2013.7588.

6. Siontis KC, Noseworthy PA, Attia ZI, Friedman PA. Artificial intelligence-enhanced electrocardiography in cardiovascular disease management. Nat Rev Cardiol. 2021;18(7):465–78. doi:10.1038/s41569-020-00503-2.

7. Attia ZI, Kapa S, Lopez-Jimenez F, McKie PM, Ladewig DJ, Satam G, et al. Screening for cardiac contractile dysfunction using an artificial intelligence-enabled electrocardiogram. Nat Med. 2019;25(1):70–4. doi:10.1038/s41591-018-0240-2.

8. Vaid A, Johnson KW, Badgeley MA, Somani SS, Bicak M, Landi I, et al. Using deep-learning algorithms to simultaneously identify right and left ventricular dysfunction from the electrocardiogram. JACC Cardiovasc Imaging. 2022;15(3):395–410. doi:10.1016/j.jcmg.2021.08.004.

9. Kwon JM, Kim KH, Jeon KH, Kim HM, Kim MJ, Lim SM, et al. Development and validation of deep-learning algorithm for electrocardiography-based heart failure identification. Korean Circ J. 2019;49(7):629–39. doi:10.4070/kcj.2018.0446.

10. Kashou AH, Medina-Inojosa JR, Noseworthy PA, Rodeheffer RJ, Lopez-Jimenez F, Attia ZI, et al. Artificial intelligence-augmented electrocardiogram detection of left ventricular systolic dysfunction in the general population. Mayo Clin Proc. 2021;96(10):2576–86. doi:10.1016/j.mayocp.2021.02.029.

11. Yao X, Rushlow DR, Inselman JW, McCoy RG, Thacher TD, Behnken EM, et al. Artificial intelligence-enabled electrocardiograms for identification of patients with low ejection fraction: a pragmatic, randomized clinical trial. Nat Med. 2021;27(5):815–9. doi:10.1038/s41591-021-01335-4.

12. Yagi R, Goto S, Katsumata Y, MacRae CA, Deo RC. Importance of external validation and subgroup analysis of artificial intelligence in the detection of low ejection fraction from electrocardiograms. Eur Heart J Digit Health. 2022;3(4):654–7. doi:10.1093/ehjdh/ztac065.

13. Carter RE, Johnson PW, Strom JB, Waks JW, Krumerman A, Ferrick KJ, et al. Multisite, external validation of an AI-enabled ECG algorithm for detection of low ejection fraction. JACC Adv. 2026;5(2):102537. doi:10.1016/j.jacadv.2025.102537.

14. Gow B, Pollard T, Nathanson LA, Johnson A, Moody B, Fernandes C, et al. MIMIC-IV-ECG: diagnostic electrocardiogram matched subset (version 1.0) [dataset]. PhysioNet. 2023. doi:10.13026/4nqg-sb35.

15. Sangha V, Khunte A, Holste G, Mortazavi BJ, Wang Z, Oikonomou EK, et al. Biometric contrastive learning for data-efficient deep learning from electrocardiographic images. J Am Med Inform Assoc. 2024;31(4):855–65. doi:10.1093/jamia/ocae002.

16. Mehari T, Strodthoff N. Towards quantitative precision for ECG analysis: leveraging state space models, self-supervision and patient metadata. IEEE J Biomed Health Inform. 2023;27(11):5326–34. doi:10.1109/JBHI.2023.3310989.

17. Noseworthy PA, Attia ZI, Brewer LC, Hayes SN, Yao X, Kapa S, et al. Assessing and mitigating bias in medical artificial intelligence: the effects of race and ethnicity on a deep learning model for ECG analysis. Circ Arrhythm Electrophysiol. 2020;13(3):e007988. doi:10.1161/CIRCEP.119.007988.

18. Bozkurt B, Coats AJ, Tsutsui H, Abdelhamid M, Adamopoulos S, Albert N, et al. Universal definition and classification of heart failure: a report of the Heart Failure Society of America, Heart Failure Association of the European Society of Cardiology, Japanese Heart Failure Society and Writing Committee of the Universal Definition of Heart Failure. J Card Fail. 2021;27(4):387–413. doi:10.1016/j.cardfail.2021.01.022.

19. Gu A, Dao T. Mamba: linear-time sequence modeling with selective state spaces. arXiv:2312.00752 [Preprint]. 2023. doi:10.48550/arXiv.2312.00752.

20. Mehari T, Strodthoff N. Advancing the state-of-the-art for ECG analysis through structured state space models. arXiv:2211.07579 [Preprint]. 2022. doi:10.48550/arXiv.2211.07579.

21. Qiang Y, Dong X, Liu X, Yang Y, Fang Y, Dou J. ECGMamba: towards efficient ECG classification with BiSSM. arXiv:2406.10098 [Preprint]. 2024. doi:10.48550/arXiv.2406.10098.

22. Zhu L, Liao B, Zhang Q, Wang X, Liu W, Wang X. Vision Mamba: efficient visual representation learning with bidirectional state space model. In: Proceedings of the 41st International Conference on Machine Learning (ICML); 2024. arXiv:2401.09417. doi:10.48550/arXiv.2401.09417.

23. Assran M, Duval Q, Misra I, Bojanowski P, Vincent P, Rabbat M, et al. Self-supervised learning from images with a joint-embedding predictive architecture. In: Proceedings of the IEEE/CVF Conference on Computer Vision and Pattern Recognition (CVPR); 2023. p. 15619-29. arXiv:2301.08243. doi:10.1109/CVPR52729.2023.01499.

24. Bossuyt PM, Reitsma JB, Bruns DE, Gatsonis CA, Glasziou PP, Irwig L, et al. STARD 2015: an updated list of essential items for reporting diagnostic accuracy studies. BMJ. 2015;351:h5527. doi:10.1136/bmj.h5527.

25. Bachtiger P, Petri CF, Scott FE, Ri Park S, Kelshiker MA, Sahemey HK, et al. Point-of-care screening for heart failure with reduced ejection fraction using artificial intelligence during ECG-enabled stethoscope examination in London, UK: a prospective, observational, multicentre study. Lancet Digit Health. 2022;4(2):e117–25. doi:10.1016/S2589-7500(21)00256-9.

26. Attia ZI, Harmon DM, Dugan J, Manka L, Lopez-Jimenez F, Lerman A, et al. Prospective evaluation of smartwatch-enabled detection of left ventricular dysfunction. Nat Med. 2022;28(12):2497–503. doi:10.1038/s41591-022-02053-1.

27. Vaid A, Jiang J, Sawant A, Lerakis S, Argulian E, Ahuja Y, et al. A foundational vision transformer improves diagnostic performance for electrocardiograms. NPJ Digit Med. 2023;6(1):108. doi:10.1038/s41746-023-00840-9.

28. McKeen K, Masood S, Toma A, Rubin B, Wang B. ECG-FM: an open electrocardiogram foundation model. JAMIA Open. 2025;8(5):ooaf122. doi:10.1093/jamiaopen/ooaf122.

29. Yang Y, Peracchio L, Mayourian J, Miller T, La Cava WG. An ECG foundation model for generalizable cardiac function prediction across the lifespan. medRxiv [Preprint]. 2026. doi:10.64898/2026.05.26.26354128.

30. Lang RM, Badano LP, Mor-Avi V, Afilalo J, Armstrong A, Ernande L, et al. Recommendations for cardiac chamber quantification by echocardiography in adults: an update from the American Society of Echocardiography and the European Association of Cardiovascular Imaging. J Am Soc Echocardiogr. 2015;28(1):1–39.e14. doi:10.1016/j.echo.2014.10.003.

